# Can long-term COVID-19 vaccination be improved by serological surveillance?: a modeling study for Mozambique

**DOI:** 10.1101/2023.08.29.23294793

**Authors:** Carol Y Liu, Kayoko Shioda, Alicia NM Kraay, Sergio Massora, Áuria de Jesus, Arsénia Massinga, Celso Monjane, Saad B Omer, Samuel M Jenness, Kristin Nelson, Stefan Flasche, Inacio Mandomando, Benjamin A Lopman

**Affiliations:** Department of Epidemiology, Emory University Rollins School of Public Health, Atlanta, GA, USA; Department of Global Health, Boston University School of Public Health, Boston, MA, USA; Department of Kinesiology and Community Health, College of Applied Health Sciences, University of Illinois Urbana-Champaign, IL, USA; Institute for Genomic Biology, Unviersity of Illinois Urbana-Champaign, IL, USA; Centro de Investigação em Saúde de Manhiça (CISM), Maputo CP1929; Mozambique Instituto Nacional de Saúde, Maputo, Mozambique; School of Medicine, Yale University; Centre for Mathematical Modelling of Infectious Diseases, London School of Hygiene and Tropical Medicine

## Abstract

Seroprevalence provides an estimate of the population-level susceptibility to infection. In this study, we used a transmission model to examine the potential of using serological surveillance to inform the timing of COVID-19 boosters in Mozambique. We simulated using population-level seroprevalence thresholds as an estimate of the risk of outbreaks to trigger the timing of re-vaccination campaigns among older adults. We compare this approach to a strategy of re-vaccination at fixed time intervals. Vaccinating older adults each time the seroprevalence among older adults falls below 50% and 80% resulted in medians of 20% and 71% reduction in deaths, respectively, and number-needed-to-vaccinate to avert one death (NNT) of 1,499 (2.5^th^-97.5^th^ centile:1,252-1,905) and 3,151 (2,943-3,429), respectively. In comparison, biennial and annual re-vaccination of older adults resulted in medians of 35% and 52% deaths averted, respectively, and NNTs of 1,443 (1,223-1,733) and 1,941 (1,805-2,112), respectively. We conducted sensitivity analysis over a range of antibody waning rates and epidemic scenarios and found that re-vaccination trigger thresholds of 50-60% seroprevalence are most likely to be efficient compared to fixed-time strategies. However, given marginal gains in efficiency even in the best-case scenarios, our results favor the use of simpler fixed-time strategies for long-term control of SARS-CoV-2.

## Background

Vaccines are a pivotal tool for SARS-CoV-2 control, eliciting strong protection against severe disease and death for vaccinated individuals^1–3^. Since the beginning of global vaccine rollout in December 2020, COVID-19 vaccines averted an estimated 19.8 million deaths in 185 countries and territories^4^. Vaccination programs enabled safer relaxation of non-pharmaceutical interventions (e.g., social distancing), which facilitated a transition away from the most severe phase of the pandemic^5,6^.

Despite early successes of SARS-CoV-2 vaccines, evidence of waning immunity and the emergence of novel immune-escaping variants raised concern over the longevity of vaccine-induced protection. Since the rise of the highly infectious Omicron variant, effectiveness of the primary series of mRNA vaccines against hospitalizations reduced by almost half compared to protection estimated from the first clinical trials^7^. Booster doses partially restored short-term protection, prompting their strong recommendation for those most at risk of developing severe outcomes^8,9^. Repeated booster campaigns aimed at restoring protection against severe disease among high risk groups remains an important tool in the medium-to long-term^10,11^.

Effective deployment of vaccines maximizes their public health impact. While population immunity against SARS-CoV-2 was still low, targeted vaccine prioritization rapidly increased protection for vulnerable portions of the population, beginning with those at the highest risk for severe outcomes and deaths. Long-term control of COVID-19 requires the consideration of refined and information-driven strategies that optimizes vaccine efficiency, ideally preventing the greatest number of severe health outcomes with the fewest resources. Identifying critical time periods and targeting population groups most susceptible to impending waves can minimize resource needs while maximizing public health impact.

Serology, a marker for prior exposure determined by the presence of antibodies against SARS-CoV-2 in blood serum, also provides information on the degree of susceptibility to infection or disease in individuals. Seroprevalence of randomly sampled individuals provides an estimation of population-level susceptibility, and has previously influenced vaccination strategy. When serological studies from England^12,13^ identified waning SARS-CoV-2 seroprevalence among the oldest age groups, an additional booster targeting this group was recommended. Seroprevalence estimates are further leveraged to guide vaccination strategies for endemic infections. In the case of measles, a fully-immunizing infection with a high reproduction number, seroprevalence is used to identify locations and age groups with inadequate immunity for targeted vaccination campaigns. By providing snapshots of the landscape of population-level immunity before surges in hospitalizations and deaths, seroprevalence estimates enabled preemptive vaccination of the most susceptible population groups^14^.

The use of serological surveillance, specifically measuring seroprevalence, to monitor changing immunity for SARS-CoV-2 at the population level and to trigger vaccination campaigns emerged as a potential long-term strategy for COVID-19 control^15,16^. However, the utility of a long-term serology-guided vaccination strategy for a pathogen with an imperfect correlate of protection is unknown and will likely depend on unpredictable long-term dynamics of SARS-CoV-2 driven by waning immunity and new variant emergence^17,18^. Targeted vaccination strategies hold potential in resource limited settings where vaccine provision is constrained^19,20^; however, there are few mathematical models tailored to localized and distinct epidemic patterns in low income countries despite their utility for optimizing vaccine strategies.

Our study uses a mathematical model of SARS-CoV-2 to determine the utility of incorporating seroprevalence to trigger future COVID-19 re-vaccination efforts. We developed our model to represent Mozambique, a resource-limited setting. Mozambique is notable for its early efforts in measuring countrywide SARS-CoV-2 seroprevalence^21^ and for studying seroprevalence in children before commencing a resource-intensive campaign to vaccinate children^22^. We assess the efficiency of a serology-guided re-vaccination strategy for COVID-19 in Mozambique under uncertain epidemic dynamics over a 10-year time horizon.

## Results

We modeled transmission dynamics and re-vaccination scenarios using a deterministic, compartmental SEIR-like model^23,24^ over a ten-year period starting in September 2022. The model was stratified by age group (≤18 years, 19-49 and ≥50 years), urban/rural and 12 immunological tiers: combinations of four levels of vaccination status (unvaccinated to three doses) and three levels of SARS-CoV-2 exposure status (unexposed to two prior exposures) with lower susceptibility for increased exposure. For long-term SARS-CoV-2 dynamics, we incorporated seasonality^25,26^ assuming annual increases in transmission in the cool, dry season (April to July) and uncertainty in transmission intensity (annual R_t_). We assessed a strategy of repeat boosting for older adults (≥50 years), likely the most cost-effective^27–30^, and compared the impact of timing additional boosters guided by population-level seroprevalence estimates to the impact of vaccinating at fixed time intervals. We integrated empirical data from Mozambique on vaccine coverage, human contact patterns collected during the pandemic and survey-derived seroprevalence.

### Model calibration

Our historical model projection matched the COVID-19 epidemic waves observed in Mozambique between 2020 and August 2022 (Fig 1) and our modeled seroprevalence closely reflected historical estimates sampled at various times points in Mozambique across three age groups in both urban and rural settings (Fig 1 & SI.7).

**Figure 1.**
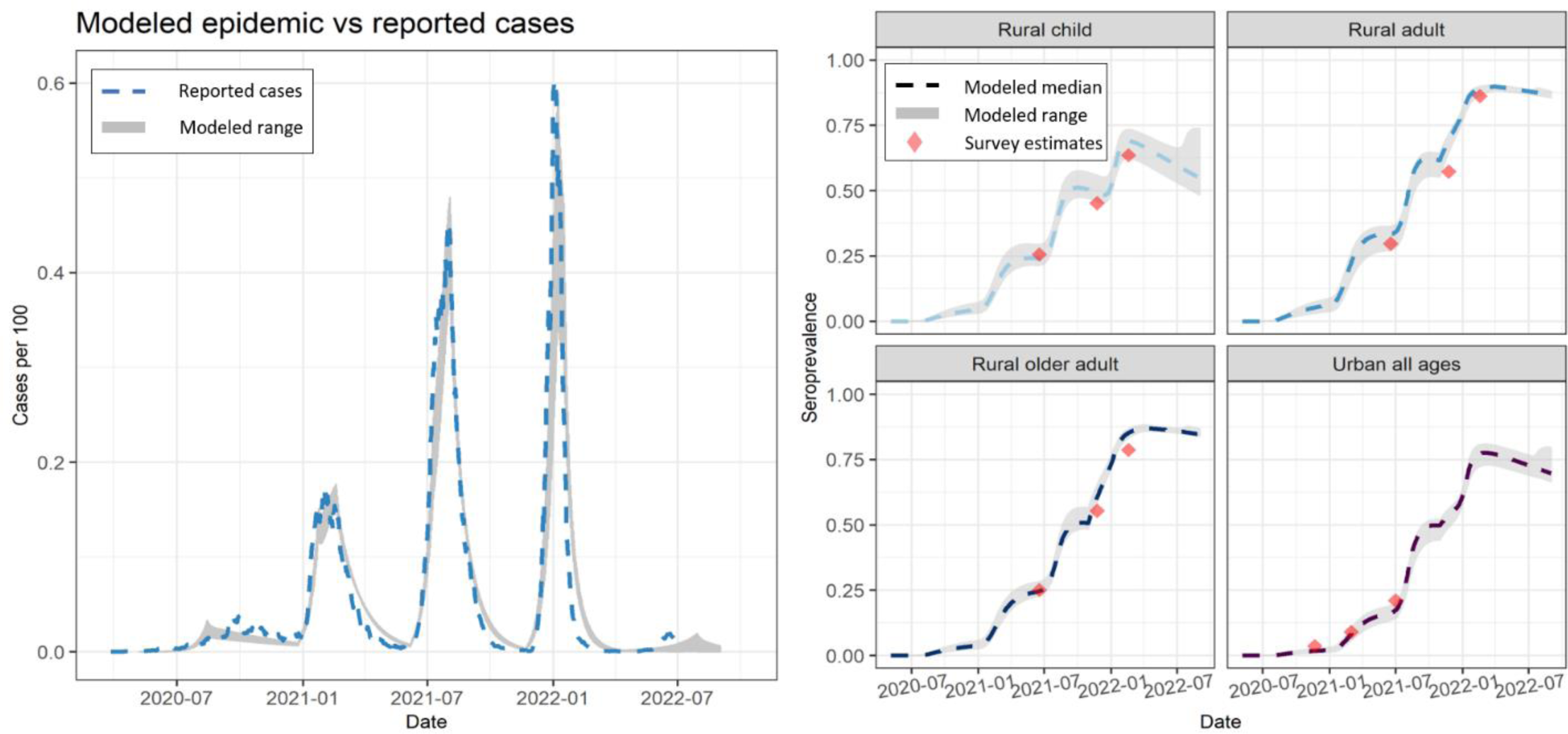
(Left) Modeled historical epidemic of cases per 100 persons using the top performing calibrated parameters (shaded grey=range of modeled cases per 100, dotted blue=reported cases per 100 accounting for 1/90 underreporting for the first two waves, 1/100 underreporting for the third wave, 1/120 underreporting for the fourth wave). Ranges reflect choice of calibration rather than uncertainty (Right) Modeled seroprevalence compared to seroprevalence estimated from serological studies during various periods of the pandemic in Mozambique.

At the start of model projections (September 2022), the median seroprevalence was 56.5%, 86.0% and 84.3% for children, adults, older adults, respectively, based on model calibration. At this time, the estimated distribution of individuals who were fully susceptible (no exposure through infection or vaccination), partially susceptible and fully immune was 16.6%, 67.0%, 16.4% for children, 0.6%, 67.9%, 31.5% for adults and 1.3%,68.3%, 30.6% for older adults. Calibrated primary series vaccine coverage among adults and older adults was 93% and 96%, respectively, comparable to the data. For children who were ineligible for vaccination, immunity was acquired from natural infection alone.

### Description of simulated epidemic and changing immunity

Long-term COVID-19 epidemic patterns will likely be driven by immune waning and immune escape^17,31^. We focused on modelling long-term dynamics driven by the former, assuming that a re-vaccination strategy triggered by seroprevalence is more likely to be beneficial under an epidemic driven by waning immunity. Under this assumption, the ten-year projection with no re-vaccination results in multiple peaks that are on average most intense in the second year and diminish over time due to an increase in population-level immunity (Fig 2). The median cumulative number of deaths across sampled R_t_ (mean = 5.5, SD = 0.2) over 10 years is 8605 (2.5th-97.5th percentile: 8026-9218) for all ages, 100 (93-108) for children, 2224 (2066-2387) for adults and 6281 (5860-6722) for older adults (SI.5.1). Seroprevalence declines over time and increases modestly following surges in cases.

**Figure 2.**
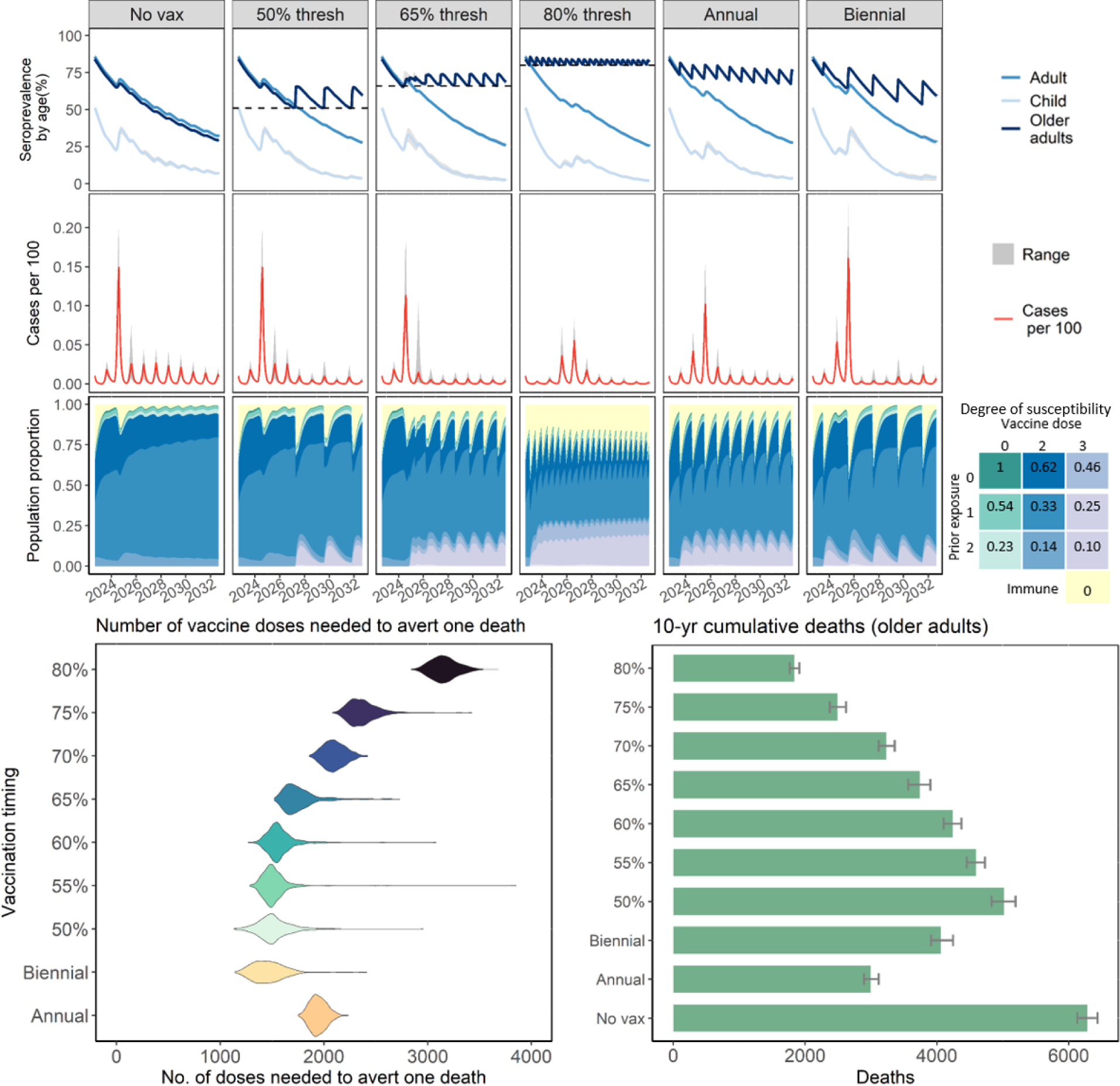
(Top row) Modeled 10-year seroprevalence (gray = 2.5^th^-97.5^th^ percentile) over time for children (lightest blue), adults (medium blue) and older adults >50 years (darkest blue) under 1) no additional vaccinations; 2) re-vaccinations timed by seroprevalence trigger thresholds of 50%, 65% and 80% and 3) re-vaccinations timed annually and biennially. (Middle row) Modeled 10-year cases per 100 individuals over re-vaccination scenarios (gray shading=ranges from random R_t_ sampling, red= median); (Bottom row) Susceptibility among older adults. Colored density represents population proportion within susceptible or immune tiers over time, ranging from fully immune (yellow) to up to 2 prior infections and 3 vaccination doses. The blue and purple densities indicate proportion of individuals in the 2- and 3-vaccine dose susceptibility tiers, respectively. The darkest shades within each color have no prior infection and are the most susceptible, with lighter shades indicating more exposure and decreased susceptibility. Individuals can wane from the 3-dose susceptibility tier to the 2-dose tier and from the 3-prior infection tier to the 2-prior infections at a rate of 1/365 days. The degree of susceptibility is indicated in the grid legend and is relative to totally susceptible, with 1 indicating fully susceptible and 0 fully immune. In scenarios with re-vaccination, campaigns generate spikes in the proportion of individuals fully immune (yellow). More frequent vaccination campaigns result in higher proportion immune (yellow) and longer period in the compartment with highest protection (purple). (Bottom left) Distribution of the number of vaccine doses needed to avert one death (NNT) by re-vaccination timing strategy (biennial, annual, triggered based on seroprevalence thresholds of between 50%-80%); (Bottom right) Cumulative deaths over ten years among older adults. Error bar represents 2.5^th^-97.5^th^ percentile of cumulative deaths for older adults.

We further observed from our model that 31% of older adults were fully immune before the start of the forward simulation, of which 58% had protection from vaccination alone with no prior infection. The proportion immune reduces over the next year as individuals lose their full protection and become increasingly susceptible to infection. This immunity gap drives a large wave in 2025 which infects 30% of the population, resulting in a shift in susceptibility whereby individuals transition from vaccine-only protection to hybrid protection from both vaccination and infection (Fig 2). The shift leads to smaller subsequent waves, in line with evidence of increased protection from hybrid immunity^32–35^.

### Descriptive results from re-vaccination strategies

For all re-vaccination scenarios, we trigger a vaccination campaign where 50% of the older adult population is vaccinated over a 30-day period. A strategy in which re-vaccination is triggered at a higher seroprevalence threshold (ex. 80%) increases the number of vaccination campaigns and cumulative vaccine doses needed. While only three campaigns (or 1.9 million doses) are needed to maintain a seroprevalence of at least 50% among older adults, 22 campaigns (or 13.9 million doses) would be needed to maintain a seroprevalence of at least 80% (SI.5.1). Using a 50% seroprevalence threshold, re-vaccination is first triggered after 4.5 years. Given the higher intensity of earlier epidemic waves, the late re-vaccination trigger has little impact on the epidemic trajectory and total disease burden. In comparison, using a 65% and 80% seroprevalence threshold triggers the first re-vaccinations after 2.3 years and 109 days, respectively. Using either of these thresholds to trigger re-vaccination reduces the size of early waves compared to the 50% threshold, but is at the expense of more frequent vaccination campaigns. In the fixed annual and biennial re-vaccination strategies implemented without regard to seroprevalence, we simulated the first re-vaccinations at 300 days for a total of 10 and 5 campaigns over ten years, respectively, early enough to reduce the size of larger projected epidemic waves in 2025.

### Impact of different re-vaccination strategy on vaccine efficiency

Compared to a median of 6,281 total simulated deaths among older adults with no additional vaccinations over ten years, vaccinating older adults each time the seroprevalence among older adults falls below 50% and 80% results in a median of 5,017 (2.5^th^-97.5^th^ percentile: 4,504-5,507) and 1,835 (1,602-2,042) deaths, respectively (Fig.2 and SI.5.1), a reduction of 20 and71%. The number needed to vaccinate to avert one death (NNT) reaches a minimum at a 50% threshold where 3 campaigns result in a median of 1,699 (1,445-1,875) fewer deaths and a median NNT of 1,499 (1,252-1,905). The NNT increases with increasing seroprevalence threshold with a median NNT of 3,151 (2,943-3,429) for an 80% threshold. In comparison, annual and biennial re-vaccination of older adults results in a median of 2,999 (2,703-3,315) and 4,059 (3,646-4,579) deaths respectively and median NNTs of 1,941 (1,805-2,112) and 1,443 (1,223-1,733), respectively (Fig.2 and SI.5.1). In summary, vaccinating at seroprevalence thresholds of 50% - 65% is more efficient than an annual strategy but less efficient than a biennial strategy.

### Tradeoffs in number-needed-to-treat

We explore two tradeoffs in the NNT of different re-vaccination strategies: tradeoffs in 1) NNT over time; 2) NNT and the number of deaths. The cumulative NNT is calculated by dividing the cumulative number of vaccine doses by the cumulative number of deaths averted by the end of year. (Fig 3). The cumulative NNT is sensitive to the timing of the first vaccination, but converges in later years. For example, using the 50% serological threshold delays vaccination until year 4 resulting in a high NNT in year 4 compared to fixed-time strategies. However, despite an early disadvantage in efficiency due to delayed vaccination, the 50% threshold achieves parity with fixed-time strategies which first vaccinate in the year 1 for maximum impact. Across all re-vaccination strategies, the NNT rises over time suggesting that re-vaccination strategies become less efficient as epidemic waves reduce.

**Figure 3.**
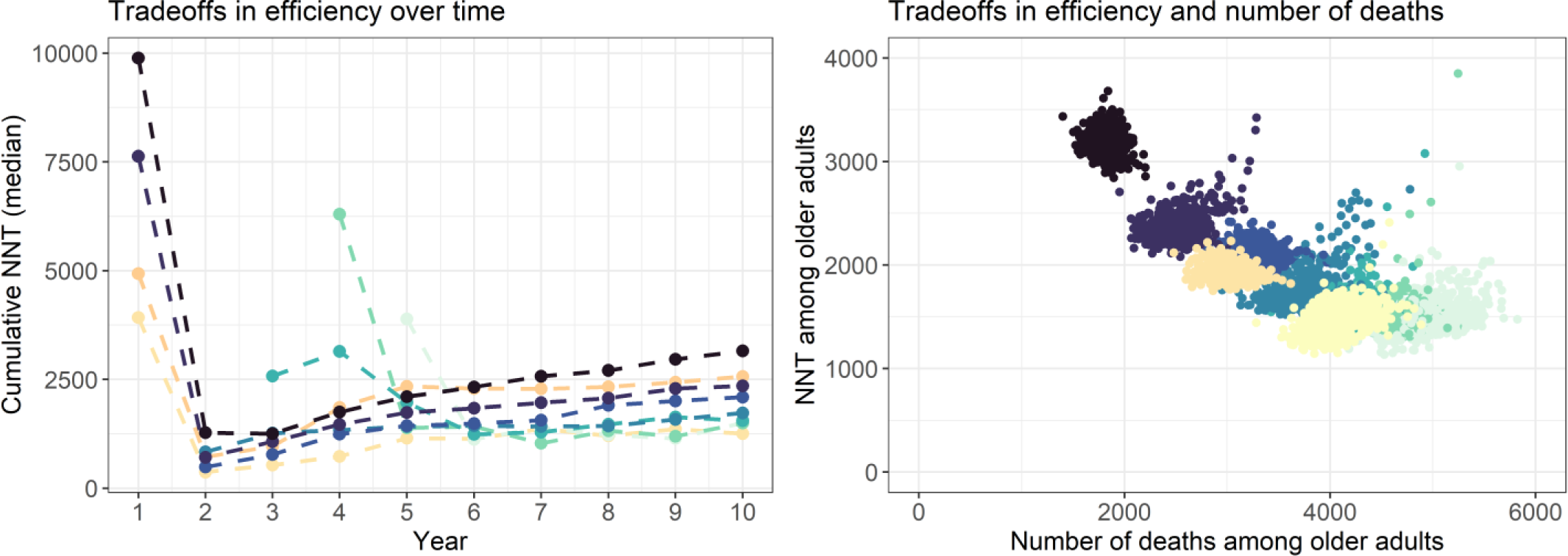
(Left) Tradeoffs over time in efficiency estimated by the cumulative number of vaccine doses needed to avert one death (NNT) for the high waning immunity epidemic scenario. Cumulative NNT is calculated by summing the number of vaccine doses provide by the end of each simulation year and dividing by the cumulative number of deaths averted by the end of each year. (Right) Scatterplot of tradeoffs in efficiency (NNT) and number of deaths for the high waning immunity epidemic scenario.

**Figure 4.**
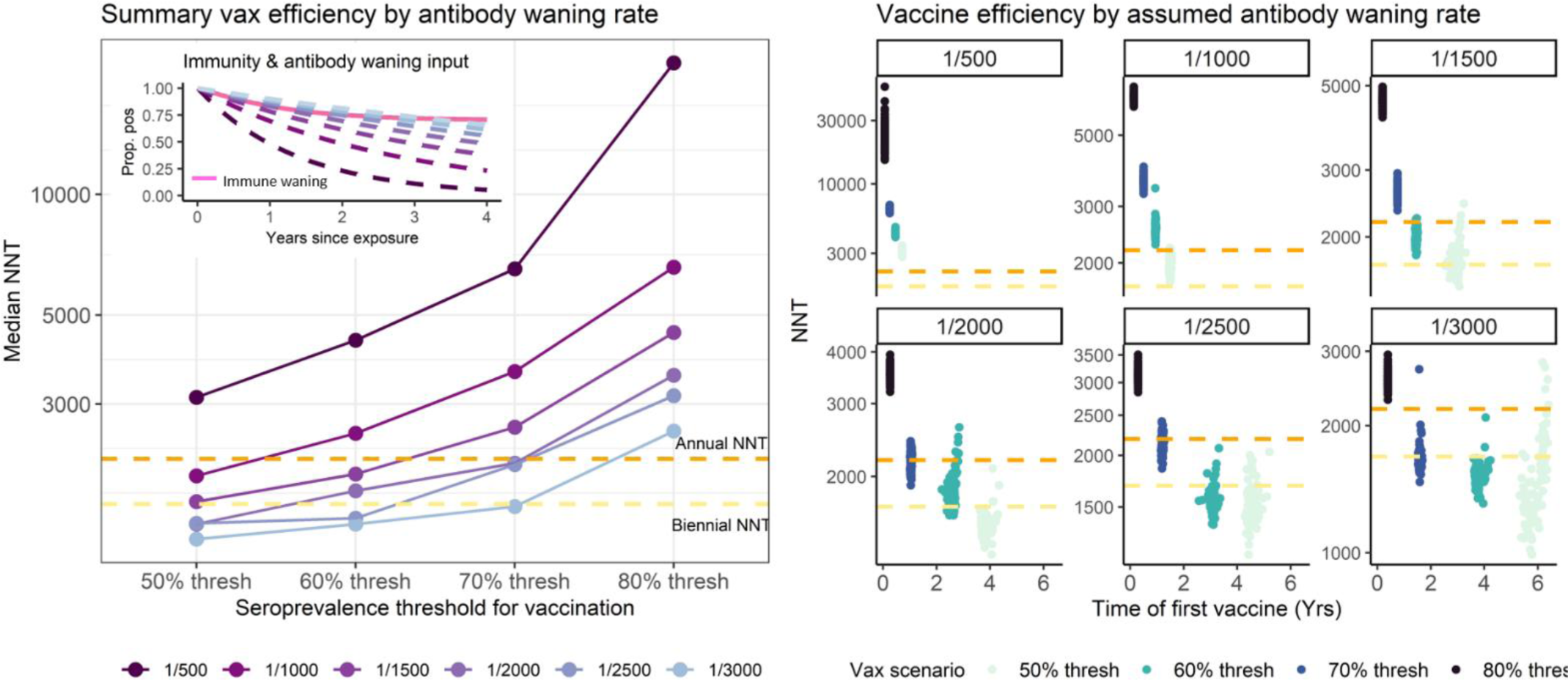
(Left) Median NNT by seroprevalence threshold for re-vaccination over various rates of time-to-seroreversion (lightest purple = slowest time-to-seroreversion, darkest purple = fastest time-to-seroreversion), with annual (orange) and biennial (yellow) NNTs for reference. Inset shows the modeled rates of different antibody waning compared to waning immunity (pink). (Right) Scatter plot of vaccine efficiency (NNT) and time of first vaccination over random Rt simulation runs under different rates of time-to-seroreversion.

We further observe a pattern of tradeoff where no single re-vaccination strategy can minimize both NNT and the cumulative deaths. Vaccinating at 80% seroprevalence minimizes the number of deaths but has a higher NNT compared to vaccinating at 50% seroprevalence and thus less efficient. Biennial re-vaccinations and re-vaccinating at 50% and 55% seroprevalence minimizes the NNT. Across these three strategies, the biennial strategy produces the lowest number of deaths compared to the two serology-triggered strategies, largely owing to an earlier timing of first vaccination campaign (Fig 3).

### Sensitivity analysis of waning antibody rates

The time-to-seroreversion of serological markers can vary by target and the choice of titer thresholds used to determine seropostivity^36^. For example, estimates of time-to-seroreversion after one infection exposure range from 250 days to 730 days for anti-N IgG and 255 days to 1500 days for anti-S IgG^36–38^, with evidence suggesting more durable antibodies after repeated exposures^13,39,40^. We vary the modeled rate of antibody waning to explore outcomes across a range of antibody targets and seroprevalence trigger thresholds that could be used as part of a surveillance program, assuming multiple exposures, from fastest (time to seroreversion of 500 days) to slowest (time to seroreversion of 3000 days). Under assumptions of relatively durable hybrid immunity after multiple exposures, we find that choosing a serological marker with rapid time-to-seroreversion relative to waning immunity results in early first vaccination and frequent subsequent re-vaccination, a highly inefficient strategy. A marker with slower waning and higher correlation with waning immunity is more likely to be efficient compared to annual and biennial re-vaccination strategies.

### Sensitivity analysis on epidemic assumptions

While the expectation is that, SARS-CoV-2 will begin to display regular, seasonal epidemic patterns, so far, SARS-CoV-2 waves have occurred at irregular times throughout the year^41^, which may continue into the near future. We conducted sensitivity analysis to explore the relative vaccine efficiency of serologically-triggered versus fixed-time re-vaccination strategies under randomly-timed epidemics by randomizing the timing of annual increases in transmission each year. We find that triggering re-vaccination at the 50% seroprevalence threshold is more efficient than both annual and biennial re-vaccinations. Compared to seasonal epidemic patterns, under randomly-timed epidemics lower trigger thresholds for serologically-guided scenarios displayed wide variations in efficiency and deaths averted. In some simulation runs, serologically-triggered strategies can be highly efficient while in others, sustained transmission over several years delayed the timing of first vaccination leading to almost no deaths averted (SI.6).

In a separate sensitivity analysis, we simulate future waves driven by increasingly transmissible immune escape variants. We find a higher number of deaths, fewer deaths averted by re-vaccination and lower efficiency but similar patterns in efficiency across re-vaccination timing strategies whereby serologically-triggered strategies are not substantially more efficient than either annual or biennial strategies (SI.7).

## Discussion

We use a transmission model to examine the potential for serological surveillance to guide the timing of future rounds of COVID-19 re-vaccination in Mozambique, a resource-limited setting. The effectiveness of timing vaccination campaigns guided by population-level prevalence thresholds of a serological marker will depend on future SARS-CoV-2 epidemic patterns and the degree to which the serological marker indicates population-level immune status. Across scenarios of waning rates of serological marker and epidemic patterns we explored, only a lower seroprevalence threshold for triggering re-vaccination (50-60%) is likely to outperform a fixed-time re-vaccination strategy with respect to efficiency. However, routine population-based sampling that can yield accurate and timely seroprevalence estimates will be costly. Given minimal gains in efficiency even in the best-case scenarios, we conclude that there is unlikely to be a cost-effective way to monitor population-level protection and reactively vaccinate the most vulnerable population groups before observing increases in clinical cases and hospitalizations. Further, when weighing tradeoffs between efficiency and deaths, we find that the biennial strategy is most likely to maximize efficiency while minimizing deaths. Taken together, the results from our modeling work favor the use of simpler fixed-time re-vaccination interval strategies over serological-triggered re-vaccination strategies.

Our results contradicted our belief that using serology-triggered re-vaccination strategies to target time periods of greater susceptibility would substantially outperform fixed-time re-vaccination strategies. In our exploration of reasons for similar efficiencies between the two strategies, we found that modeled seroprevalence did not necessarily predict deaths. We expected a negative correlation between seroprevalence and deaths where higher seroprevalence leads to lower deaths. While this correlation was observed within each year, we found an overall positive correlation over the 10-year period (SI.5.4). Large, early epidemic waves that resulted in more deaths among older and younger populations alike were driven by lower protection among children. These larger early waves occurred despite high seroprevalence among older adults, making it a weaker marker for overall epidemic size.

Our modeling work provides a framework for explicit considerations necessary for population-level serological surveillance to guide response. The degree of correlation between seroprevalence and immunity is likely to impact its utility. In the case of SARS-CoV-2, studies have explored the level of protection conferred by titers of different serological markers at the individual-level^42,43^. To formulate a feasible population-based strategy using information on correlates of protection requires its translation into measurable population-level estimates. Selecting an appropriate serological marker and a corresponding titer threshold that can reflect complex population-level susceptibility to future outbreaks and can be monitored through population surveillance will be key. Relatedly, considerations must be given to selecting the most suitable seroprevalence trigger threshold. In the case of measles, where the goal of vaccination is to eliminate infections, the proposed serologically-guided vaccination strategy uses the herd immunity thresholds as the trigger for vaccination campaigns^14^. In contrast, the primary objective of SARS-CoV-2 vaccination is to reduce severe outcomes and deaths. Without defined population-level thresholds predictive of severe outcome potential, our modeling tested a range of seroprevalence thresholds as vaccination triggers. Our analysis demonstrated, that in the context of durable hybrid immunity against severe outcomes, using a serological marker with slower waning to trigger re-vaccination is most likely to be efficient against fixed-time strategies. These considerations are further applicable to other infectious diseases that can benefit from leveraging serosurveillance to inform public health interventions^44,45^

Our analysis is the first study to assess the efficiency of serological triggers for a long-term SARS-CoV-2 strategy. Our model structure extends previously published models^26,27^ by explicitly representing complex immunity profiles of hybrid immunity. Since the duration and extent of protection conferred by prior exposures depends on whether an individual’s particular history includes infection by the virus or one or more vaccine doses^31^, an explicit representation more accurately reflects population-level susceptibility that dynamically changes in response to vaccinations, infections and waning immunity. Unlike other models, we further decouple antibody waning from immunity waning with both processes informed by available data, which more accurately reflects their varying timelines and dynamics. Our NNT values are comparable to two other modelling studies that evaluated the efficiency of booster doses in LMICs where one estimated the NNT to be around 4000 for one booster for all eligible age groups^29^ and a second estimated the NNT to be 1453 for yearly boosters among those aged 60 years and above^30^.

We acknowledge several limitations. There is considerable uncertainty around model parameters, especially for the extent of protection conferred by multiple exposures and duration of antibody waning^46^. Our parameterization reflected the observed protection conferred by prior exposures between March and January 2021, where cross protection against repeat infections was predominantly driven by c the wild-type strain and variants up to omicron^47–49^. Future variants may have greater or less cross protection against future infections than we have modeled here. We assumed a seasonally-forced long-term transmission pattern based on evidence from other respiratory illnesses in Mozambique and from other human coronaviruses^17,29,50,51^. Their remains considerable uncertainty regarding the long-term dynamics of endemic SARS-CoV-2 infection and dynamics proposed by our model served as a base case scenario that allowed for an evaluation of the merits of using a seroprevalence-guided long-term re-vaccination strategy. We tested other epidemic patterns as sensitivity analysis and found comparable results.

In conclusion, our study demonstrates that regularly-timed re-vaccinations for older adults, particularly prior to the SARS-CoV-2 season is likely to be more efficient and similarly effective as a serological-triggered re-vaccination strategy. This finding was contrary to our expectation that a serological-triggered strategy could prevent periods of enhanced population risk due to waned protection.

## Methods

### Model structure

We extend a deterministic, compartmental SEIR-like model^23,24^ to incorporate demographic strata of age group (≤18 years, 19-49 and ≥50 years) and urban/rural and twelve tiers of immunity status: combinations of four tiers of vaccine status (unvaccinated, vaccinated with one dose, two doses and three doses) and three tiers of exposure status (unexposed, one prior exposure and two prior exposures) (Fig 5). The multiple tiers of immunity allow differential susceptibility based on prior exposure from either infection or vaccination. To summarize the model design: after exposure, individuals enter a latent, non-infectious period (E), after which they progress to either infectious and asymptomatic (A) or infectious and symptomatic (I). A proportion of symptomatic individuals progress to more severe disease and are hospitalized (H). A subset of those who are hospitalized ultimately die from SARS-CoV-2 (entering the D class). All individuals who are not hospitalized recover (entering the R class) and can either be seropositive (R_p_) or seronegative (R_n_). Individuals can also be vaccinated and, if unexposed, enter the seropositive V_p_ or seronegative V_n_ classes immediately post-vaccination, if previously exposed, enter the seropositive R_p_ or seronegative R_n_ classes corresponding to their prior infection tier. The R and V classes are temporarily immune to infections; however, immunity wanes over time and individuals return to a partially susceptible class (S_p,1_ for seropositive and one prior infection and S_n,1_ for seronegative and one prior infection). The force of infection (SI.1. Eq.1) is modified by probability of infection of exposed age group (*β*_*i*_), vaccine effectiveness against infection, differential by 1-3 doses (*VEI*_*v*_), reduced susceptibility from protection from prior infection (*IP*_*e*_), increased variant transmissibility (for Delta and Omicron waves), immune escape (applied to immunity tiers with prior exposure through either infection of vaccination), age and rural/urban-specific contact rate (*χ*_*j*,*i*,*m*,*k*_) and infection density within each demographic strata. Infected individuals who are asymptomatic have reduced transmissibility (*α*_*i*_). The model diagram is described in Figure 1, equations can be found in SI.1 and details on model parameters, values, ranges and sources can be found in SI.2.

**Figure 5.**
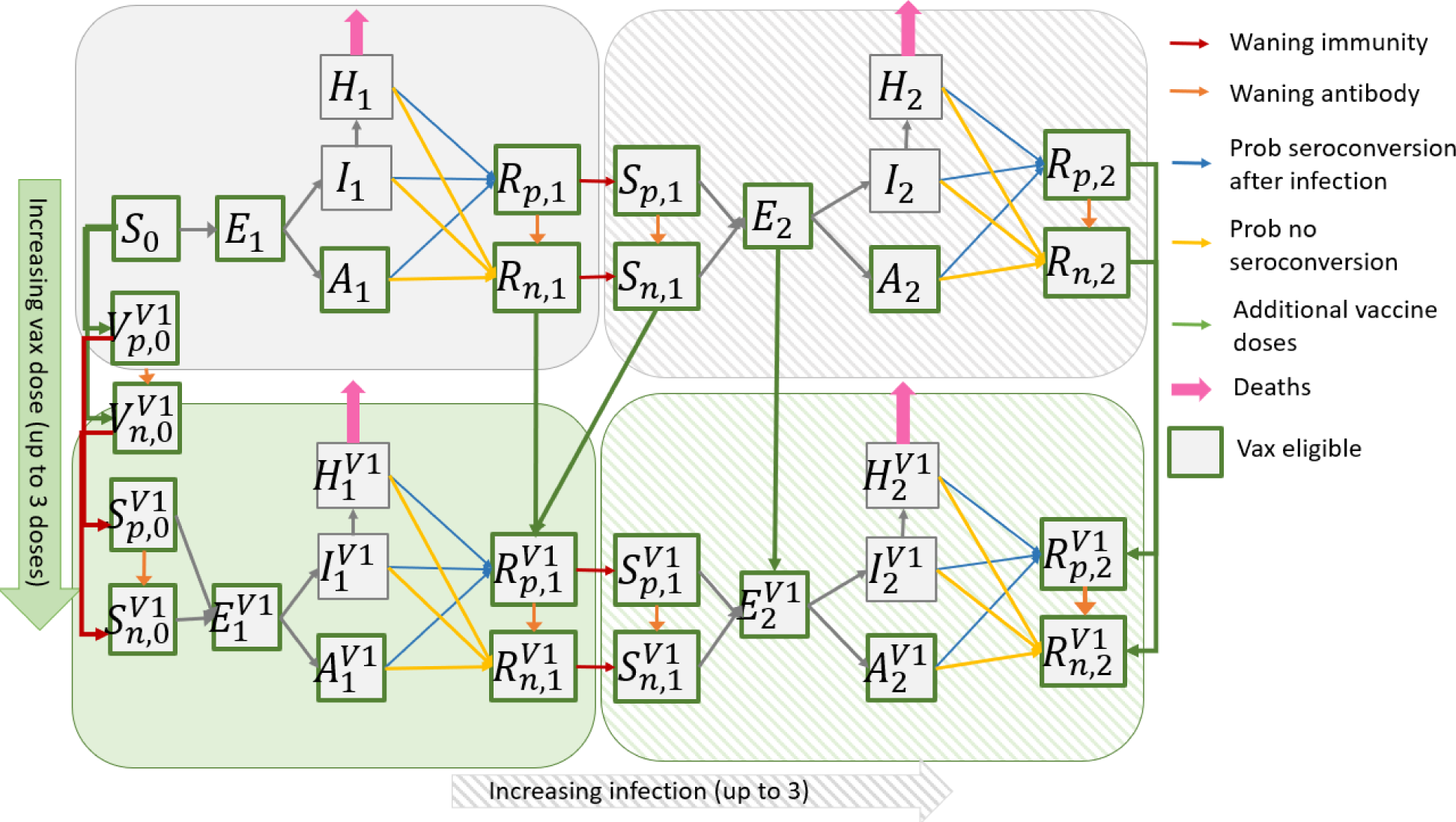
Schematic of an S-E-I-R-like compartment for a single demographic stratum (out of six total: three age groups in each of urban and rural) with four tiers of immunity shown (two tiers of vaccination: unvaccinated and one dose of vaccine, with the superscript V1 the vaccine dose and two tiers of exposure: no prior exposure and one exposure, with the subscript representing the number of exposures). Individuals who recover from infection are immune for a period. The majority seroconvert after infection (R_p_) while others do not (R_n_). Immunity for both seropositive (R_p_) and seronegative (R_n_) can wane over time, returning individuals to S_p_ and S_n_, respectively, allowing for subsequent infection. Individuals in classes outlined in green are eligible for vaccination and move to a higher vaccine tier upon vaccination (not all arrows drawn explicitly in the diagram) The majority of individuals seroconvert after vaccination. Vaccinated individuals are temporarily immune before their immunity wanes. Individuals in vaccinated and previously exposed strata have a reduced probability of infection and disease.

### Seroconversion and seroreversion

Our model distinguishes between antibody positivity and immunity with separate seropositive and seronegative compartments in the S (susceptible), R (recovered) and V (vaccinated) disease states. Presence of neutralizing antibodies following exposure is associated with reduced risk of severe disease^42,52–55^. Nevertheless, protection against infection also depends on cell-mediated immunity and circulating variants. For example, individuals who lack neutralizing antibodies but have robust cell-mediated immunity can still be protected, while others with antibodies may remain susceptible to new immune-escaping variants^42,56,57^. Explicit separation of immunity and serological status represents their imperfect correlation.

Upon exposure through either infection or vaccination, individuals can seroconvert and become seropositive. We assumed that 90%^52,53^ of infected individuals seroconvert after recovering and 85%^13^ of vaccinated individuals seroconvert upon moving to the vaccinated class following first dose. Further, we assume 70% of seronegative individuals who receive an additional vaccine dose will seroconvert^58^.

Over time, antibodies can wane with seropositive individuals seroreverting to seronegative. Fully immune and seropositive individuals (R_p_) will serorevert to fully immune and seronegative (R_p_) while partially susceptible and seropositive individuals (S_p_) will serorevert to partially susceptible and seronegative (S_n_). Waning rates differ by number of exposures, regardless of whether the exposure was from vaccination or infection (e.g. antibody waning among those infected once and vaccinated once is the same as waning among those vaccinated twice). Seroreversion is fastest following the first exposure (1/500 days)^59–62^ and declines after multiple exposures (range of 1/2500 days after second exposure), in line with recent evidence that antibody titers are higher and more persistent after multiple exposures^63,64^. To more appropriately represent the dynamics of waning antibodies in the real world, the rate of seroreversion is modeled as a gamma distribution with four sequential compartments for the tiers with the fastest waning rates^65^ (SI.1 for details).

### Tiered susceptibility

Following infection or vaccination, we assume that individuals are fully immune for an average of 150 days before moving to a partially susceptible class. Among unvaccinated individuals, one and two prior infections will confer 65%^32^ and 75% protection against infection (*IP*_*e*_), respectively.

Vaccinated individuals move to a higher vaccination class (V1 → V2 → V3) with reduced rates of infection and probability of hospitalization if infected. We parameterize vaccine effectiveness based on performance of Astra Zeneca, the main vaccine used in Mozambique (and many LMICs) and assume that one dose of vaccine reduces rate of infection (*VEI*_*v*_) by 50%, two doses by 60% and three doses by 70%. Further, one, two, three doses of vaccine reduces the probability of progression to severe disease (*VEP*_*v*_) by 40%, 67% and 70% respectively. Overall vaccine effectiveness against hospitalization is 70%, 87% and 91%^66^ for one, two, three doses of vaccine, respectively. For individuals with hybrid immunity, protection from infection is determined by 1-(1-*IP*_*e*_)*(1-*VEI*_*v*_). SI.13 and SI.14 summarizes key evidence on infection and vaccination effectiveness against susceptibility and severe disease stratified by variants used to inform our immunity parameters.

### Data sources and calibration

Model calibration provided an estimation of the population distribution across compartments and the seroprevalence at the start of the simulation (Sept 1, 2022), nine months after the last sampled seroprevalence in Mozambique. We incorporated data on social contact sampled in an urban and a rural area in Mozambique during the COVID-19 pandemic between March 2021-March 2022; multiple cross-sectional seroprevalence data available at several time points from an urban and a rural area (SI.3.3) in Mozambique; vaccination data and time-series of reported cases^67^ adjusted for an underreporting factor to calibrate the model.

The following parameters were calibrated using an approximate Bayesian approach: *β*_*c*_, *β*_*a*_, *β*_*e*,_ (probability of infection upon contact among children, adults and older adults), increased transmissibility and immune escape for Delta and Omicron variants and waning rate of antibodies (SI.2). We defined a range of plausible priors for each parameter informed by literature review and prior experience calibrating a comparable model. The initial ranges for calibration can be found in SI.7. We used Latin hypercube sampling^68^ to randomly sample from the pre-defined parameter space for each run. R_0_ from the sampled *β*s was calculated by identifying the dominant eigenvalue of the next generation matrix that incorporates both age-specific mixing patterns and the age-specific probabilities of transmission (*β*_*c*_, *β*_*a*_, *β*_*e*_)^69^. We further constrained *β*s to sampled trios that met the pre-specified R_0_. We compared the modeled age-specific seroprevalence estimates after each wave to available seroprevalence information as the primary target statistic (17 unique estimates). We conducted the calibration iteratively. Initially, 5000 iterations were sampled from the initial range of parameter spaces. We then identified parameter draws that performed in the top 10% based on the sum of square errors across all seroprevalence estimates and where each modeled data point was within 5 percentage points of estimates measured from field studies. We then restricted the ranges for each parameter and conducted another round of LHS sampling based on the new, restricted ranges. This process was repeated 4 times until, iteratively narrowing the calibration range each time. We then ranked each set of parameters by the sum of square errors for seroprevalence estimates and conducted forward simulation using the top 10% (n=500). We then chose the set of parameters that produced the median cases and epidemic trajectory over the 10-years simulation period as the primary parameter values for the forward simulations. Sensitivity analysis for the range of acceptable calibrated parameters (top 10%) to assess the degree to which uncertainty in the calibrated values would affect our forward simulation results. Our calibrated values for transmissibility of the delta and omicron variants are in line with the published literature^70,71^. Ranges from calibration reflect choice in calibration rather than uncertainty.

### Forward simulation epidemiological scenarios

We simulated the epidemic forward for ten years from September 1, 2022. Dynamics of long-term immunity were simulated by allowing waning from the highest vaccination and infection tier in addition to the modeled waning described earlier. This included: 1) waning immunity for individuals who recovered after their third infection (*R*_*p*,3_, *R*_*n*,3_) to the two-infection susceptible tier (*S*_*p*,2_, *S*_*n*,2_) after a period of immunity and 2) waning immunity for individuals from the three-vaccine-dose susceptibility tier (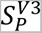, 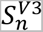) to the two-vaccine-dose susceptibility tier (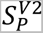, 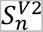) after a period of full immunity. We assumed annual waves driven by increases in transmission in the cool, dry season (April-July in Mozambique), informed by observational studies of early SARS-CoV-2 dynamics that cooler and dryer weather were moderately associated with increased SARS-CoV-2 transmissibility^25,72,73^. The relationship between *R*_0_(*t*) and the specific humidity q(t) is determined by a prior model^25^: *R*_0_(*t*) = exp(*α* ∗ *q*(*t*) + ^log(*R*^_0*max*_ − *R*_0*min*_^)) + *R*^_0*min*_.

To represent uncertainty in future transmission, we sample *R*_0*max*_ for each year from a log normal distribution (mean R_0_ = 5.5, standard deviation = 0.2). *R*_0*min*_ is fixed at 2.05, marginally lower than the transmissibility of the original Wuhan strain. Given unpredictable long-term transmission dynamics, we consider one scenario where future waves are driven by high rates of immune escape and a second scenario where future waves are driven by high rates of waning immunity. In the high immune escape scenario, in addition to sampling *R*_0*max*_ from the distribution, we enforce a general trend of yearly increases using the following formula: 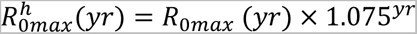, where 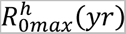 is the yearly *R*_0*max*_ for the immune escape scenario, *R*_0*max*_ (*yr*) is the randomly sampled R_0_ and *yr* is years since start of simulation. In the waning immunity scenario, we allow for additional waning immunity from the highest three-exposure tier to the two-exposure tier (90% protection to 75% protection in the three-vaccine dose tier and 86% protection to 67% protection in the two-vaccine dose tier). For each scenario, we conducted 500 different runs, each with 10 randomly sampled R_0_ for each year.

### Vaccination triggers and analytical outputs

Based on literature suggesting increased impact and cost-effectiveness of routine vaccination for older adults compared to routine vaccination of other age groups, we focus on a strategy of booster vaccination for older adults and compare the impact of timing additional doses guided by population-level seroprevalence estimates or based on fixed intervals. When triggered in the model, additional vaccination was provided to the older adult population at 2% (or ∼28,000 doses) of the older adult population per day over a 30-day campaign period for 50% coverage per campaign, with the same vaccination rate applied to seropositive and seronegative subgroups. A 30-day campaign was deemed feasible and vaccinating ∼28,000 deemed achievable compared to the 100,000 doses provided per day during peak campaign periods for the primary COVID-19 series in Mozambique^74^. Specifically, the timing of vaccination was guided by: 1) seroprevalence thresholds among older adults ranging from 50-80% where vaccination will be triggered when the seroprevalence falls below the threshold; 2) fixed time intervals where vaccination will be triggered annually or biennially, with the first vaccine campaign triggered a year after the start of simulation.

Our primary outcome is the number of vaccinations provided to older adults needed to avert one death in the population (or number-needed-to-treat, NNT), using the number of deaths when no additional vaccinations are provided as the base case. The NNT allows more equitable comparison between scenarios where vaccination is constantly triggered versus scenarios where vaccination is rarely triggered, shedding insight on the efficiency and potential cost-effectiveness of different vaccination timing strategies. We further present the number of deaths and the number of deaths averted compared to a no vaccination scenario and the number and timing of additional vaccination campaigns.

## Data Availability

All data produced in the present work are contained in the manuscript or available as part of the Github repository: https://github.com/lopmanlab/COVID_serovax_Mozambique

