## Supplementary material for "Can long-term COVID-19 vaccination be improved by serological surveillance?: a modeling study for Mozambique": SI

**Supplementary Information**

### SI.1 Model methodology

#### SI.1.1 Model equations

The first set of equations shows the compartments for one age group, urban/rural with no previous vaccination or prior exposure

$$\begin{matrix} & \frac{dS_{v=0,e=0}}{dt}=-\lambda_{v=0,e=0}\left( t \right)S_{v,e}- delta_{v1}S_{v,e} \\ & \frac{dE_{v=0,e=1}}{dt}=\lambda_{v=0,e=0}\left( t \right)S_{v,e}-\sigma E_{v,e}- delta_{v1}E_{v,e} \\ & \frac{dI_{v=0,e=1}}{dt}=\left( \nu_{i} \right)\sigma E_{v}-\gamma_{I}I_{v} \\ & \frac{dA_{v=0,e=1}}{dt}=\left( 1-\nu_{i} \right)\sigma E_{v,e}-\gamma_{A}A_{v,e}- delta_{v1}A_{v,e} \\ & \frac{dH_{v=0,e=1}}{dt}=\left( 1-VEP_{v} \right)\phi_{i}\gamma_{I}I_{v,e}-\gamma_{H}H_{v,e} \\ & \frac{dR_{v=0,e=1}^{p}}{dt}=\pi\left( 1-\left( 1-VEP_{v} \right)\phi_{i} \right)\gamma_{I}I_{v,e}+\pi\gamma_{A}A_{v,e}+\pi\left( 1-\mu_{i} \right)\gamma_{H}H_{v,e}- delta_{v1}R_{v,e}^{p}-\omega_{i}R_{v,e}^{p}-\kappa_{1}R_{v,e}^{p} \\ & \frac{dR_{v=0,e=1}^{n}}{dt}=\left( 1-\pi\right)\left( 1-\left( 1-VEP_{v} \right)\phi_{i} \right)\gamma_{I}I_{v,e}+\left( 1-\pi\right)\gamma_{A}A_{v,e}+\left( 1-\pi\right)\left( 1-\mu_{i} \right)\gamma_{H}H_{v,e}-\delta_{v1}R_{v,e}^{n}-\omega_{i}R_{v,e}^{n}+\kappa_{1}R_{v,e}^{n} \\ & \frac{dM_{v,e}}{dt}=\mu_{i}\gamma_{H}H_{v,e} \\ & \\ & \end{matrix}$$

For other demographic and immunological strata, the equations are similar except the S compartment is split into Sp and Sn for individuals who are 1) susceptible and seropositive or 2) susceptible and seronegative, respectively. Equations below show the S compartments for the immunity strata of v=1 and e=0. Note that for the tiers with the fastest antibody waning, we further implement a gamma-distributed waning for the sero-reversion process from Rp to Rn (detailed below).

$$\begin{matrix} & \frac{dS_{v=1,e=0}^{p}}{dt}=\omega_{i}V_{v,e}^{p}-\lambda_{v=1,e=0}\left( t \right)S_{v,e}^{p}-\delta_{v2}S_{v,e}^{p}-\kappa_{1}S_{v,e}^{p} \\ & \frac{dS_{v=1,e=0}^{n}}{dt}=\omega_{i}V_{v,e}^{n}-\lambda_{v=1,e=0}\left( t \right)S_{v,e}^{n}-\delta_{v2}S_{v,e}^{n}+\kappa_{1}S_{v,e}^{p} \end{matrix}$$

Likewise, equations below show the S compartments for the immunity strata of v=0 and e=1:

$$\begin{matrix} & \frac{dS_{v=0,e=1}^{p}}{dt}=\omega_{i}R_{v,e}^{p}-\lambda_{v=0,e=1}\left( t \right)S_{v,e}^{p}-\delta_{v1}S_{v,e}^{p}-{4*\kappa}_{1}S_{v,e}^{p} \\ & \frac{dS_{v=0,e=1}^{n}}{dt}=\omega_{i}R_{v,e}^{n}-\lambda_{v=0,e=1}\left( t \right)S_{v,e}^{n}-\delta_{v1}S_{v,e}^{n}+\kappa_{1}S_{v,e}^{p} \end{matrix}$$

Individuals who have no prior history of infection go into the V compartments upon vaccination with one, two or three vaccine doses, allowing a period of temporary immunity. Individuals who have a prior history of infection go into the R compartment corresponding to their prior infection tier (see model diagram in Methods section of paper). This again allows for a period of temporary immunity before returning individuals to a susceptible tier corresponding to the number of vaccine doses and number of prior infections.

$$\begin{matrix} & \frac{dV_{v=1,e=0}^{p}}{dt}=\rho_{v1} delta_{v1}S_{v,e}-\omega_{i}V_{v=1,e=0}^{p}-\kappa_{1}V_{v,e}^{p} \\ & \\ & \frac{dV_{v=1,e=0}^{n}}{dt}=\left( 1-\rho_{v1} \right) delta_{v1}S_{v,e}-\omega_{i}V_{v=1,e=0}^{n}+\kappa_{1}V_{v,e}^{n} \\ & \\ & \\ & \\ & \\ & \\ & \\ & \end{matrix}$$

The force of infection is as follows:

$$\begin{matrix} \\ \lambda_{i,k,v,e}\left( t \right)=\beta_{i}\left( 1-VEI_{v} \right)(1-{IP}_{e})(esc)(var\_trans)\left[ \sum_{j=c,a,e} \sum_{m=r,u} \left( \frac{\chi_{j,i,m,k}\sum_{e=0}^{3} \sum_{v=0}^{3} I_{j,m,v,e}\left( t \right)+\alpha A_{j,m,v,e}\left( t \right)}{\sum_{e=0}^{2} {\sum_{v=0}^{3} N_{j,m,v,e}\left( t \right)}} \right) \right] \end{matrix} (eq.1)$$

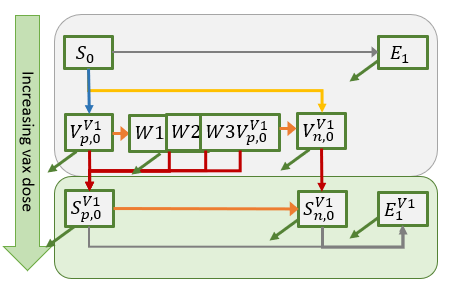

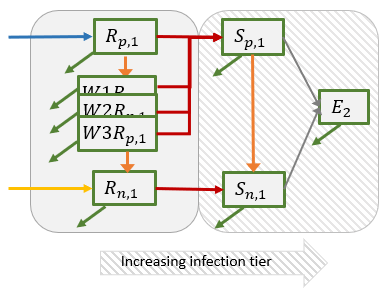
SI.1.2 Gamma-distributed antibody waning process

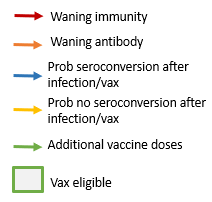

*Figure 1. Diagram describing the gamma-distributed antibody waning process for the tiers with the fastest rates of antibody waning. (Left) Gamma-distributed waning for those who were seropositive after their first infection. (Right) Gamma-distributed waning for those who were seropositive after first vaccination.*

To most appropriately represent the dynamics of antibody waning, we implement a gamma-distributed waning process for the tiers with the fastest rates of antibody waning. After first infection, individuals can be seropositive (R_p,1_) or seronegative (R_n,1_). Seropositive individuals can wane into seronegative through a gamma-distributed process with four compartments. Seropositive individuals can also lose their immunity and become susceptible again (S_p,1_) at a rate and in a process independent from the waning of antibodies. All compartments are eligible for vaccination. The equations specific to the gamma-distributed waning are detailed below. The same logic applies to the gamma-distributed waning of antibodies for those who are seropositive after receiving their first dose of vaccination.

$$\frac{dR_{v=0,e=1}^{p}}{dt}=-\omega_{i}R_{v,e}^{p}-4*\kappa_{1}R_{v,e}^{p}- delta_{v1}R_{v,e}^{p}$$

$$\frac{d{W1R}_{v=0,e=1}^{p}}{dt}=-\omega_{i}{W1R}_{v,e}^{p}-4*\kappa_{1}W1R_{v,e}^{p}+4*\kappa_{1}R_{v,e}^{p}- delta_{v1}{W1R}_{v,e}^{p}$$

$$\frac{d{W2R}_{v=0,e=1}^{p}}{dt}=-\omega_{i}{W2R}_{v,e}^{p}-4*\kappa_{1}W2R_{v,e}^{p}+4*\kappa_{1}{W1R}_{v,e}^{p}- delta_{v1}{W2R}_{v,e}^{p}$$

$$\frac{d{W3R}_{v=0,e=1}^{p}}{dt}=-\omega_{i}{W3R}_{v,e}^{p}-4*\kappa_{1}W3R_{v,e}^{p}+4*\kappa_{1}W2R_{v,e}^{p}- delta_{v1}{W3R}_{v,e}^{p}$$

$$\frac{dR_{v=0,e=1}^{n}}{dt}=4*\kappa_{1}W3R_{v,e}^{p}-\delta_{v1}R_{v,e}^{n}-\omega_{i}R_{v,e}^{n}$$

#### SI.1.3 Implementing historical vaccination

Individuals in S, E, A, R compartments are eligible for vaccination. Historical vaccinations were implemented based on documented first and second dose vaccination administered in Mozambique over time, beginning March 2021,^1^ with an early preference towards the age group of 50 years and above. At the start of the forward simulation period (Sept 1^st^, 2022), the two-dose vaccination coverage among adults is >90%, while children <18 years of age in Mozambique have not yet been vaccinated.

### SI.2 Model parameters

*Table 1****.*** *Model parameters, corresponding description, value and source*

| **Abbreviation** | **Description** | **Value (Range)** | **Source** |
| --- | --- | --- | --- |
| **Transmission** |  |  |  |
| *R0* | Basic reproduction number | 2.58 (2-4) | ^2,3^ |
| $\beta_{c},\beta_{a},\beta_{e}$ | Prob. of infection of exposed age group | 0.02326, 0.02442, 0.0116 | Calibrated |
| $\alpha$ | Relative infectiousness btwn asympt & sympt | 0.6 (0.3-0.9) | ^4,5^ |
| $\gamma_{I}$ | Infectious period, symptomatic (days) | 7 | ^6^ |
| $\gamma_{A}$ | Infectious period, asymptomatic(days) | 7 | ^6^ |
| $\gamma_{H}$ | Hospital LOS (days) | 5 (3-9) | ^7^ |
| $\sigma$ | Latent period | 5.5 (3.0-6.7) | ^8–11^ |
| $\nu_{c},\nu_{a}, \nu_{e}$ | Probability of symptomatic infection by age group | 0.45, 0.55, 0.65 | ^4^ |
| $\phi_{c}, \phi_{a}, \phi_{e},$ | Probability of hospitalization upon symptomatic infection by age group | 0.004-0.0075,  0.03-0.15,  0.2-0.35 | ^12–14^ |
| $\mu_{c}, \mu_{a}, \mu_{e}$ | Probability of death upon hospitalization by age group | 0.005-0.01,  0.0365-0.465,  0.15 | ^15,16^ |
| $\omega_{c}, \omega_{a}, \omega_{e}$ | Duration of full immunity by age group | 1/150, 1/150, 1/150 | ^17^ |
| **Serology** | | |  |
| $\pi$ | Probability of seroconversion after infection | 0.9 (0.8-0.98) | ^18,19^ |
| $\rho_{v1},\rho_{v2},\rho_{v3}$ | Probability of seroconversion among seronegative individuals after vaccination (dose 1-3) | 0.85,0.7, 0.9 | ^20^ |
| $\kappa_{1}$ | Seroreversion rate for first exposure (either vaccine or infection) | 1/500 | ^21,22^ |
| $\kappa_{2}$ | Seroreversion rate for second or more exposures exposure | 1/2500 | ^21,22^ |
| **Vaccination** | | |  |
| $\delta_{i,k,v}$ | Per capita vaccination rate by age group, urban/rural and dose | Time-varying based on data (0.01%-4%) | ^1^ |
| $VEI_{v1},VEI_{v2}, VEI_{v3}$ | Vaccine effectiveness against infections by number of vaccine doses (assuming Astra Zeneca) | 0.5, 0.6, 0.7 | ^23^ |
| $VEP_{v1},VEP_{v2}, VEP_{v3}$ | Vaccine effectiveness against progression from infection to severe disease/hospitalization by number of vaccine dose (assuming Astra Zeneca) | 0.4, 0.67, 0.9 | ^23^ |
| **Prior exposure protection** | | |  |
| ${IP}_{e}$ | Protection from first/second infection | 0.65, 0.75 | ^24^ |
| **Variants** | | |  |
| *R0 (delta)/var_trans* | R0 during delta wave/increased transmissibility during delta wave | 3.1 (2.4-4) | Calibrated^25,26^ |
| *R0 (omicron)/var_trans* | R0 during omicron/increased transmissibility during omicron | 6.4 (5.0-6.5) | Calibrated^27^ |
| *Immune escape (delta)* | Increased transmission among those with prior exposure during delta wave | 1.2 | ^28,29^ |
| *Immune escape (omicron)* | Increased transmission among those with prior exposure during omicron wave | 1.6 | ^28,29^ |
| *Immune escape (new variant)* | Increased transmission among those with prior exposure for new variant | 1.7 | Assumption |
| **Under-reporting** |  |  |  |
| *Under-reporting (wave alpha-beta)* | Case under-reporting for alpha/beta wave |  | Calibrated |
| *Under-reporting (delta)* | Case under-reporting for delta wave |  | Calibrated |
| *Under-reporting (omicron)* | Case under-reporting for omicron wave |  | Calibrated |

### SI.3 Data sources from Mozambique

#### SI.3.1 Summary of data sources on contact, seroprevalence, vaccination and cases

| Parameter | Source | Stratification |
| --- | --- | --- |
| Social contact mixing matrix | GlobalMix Study (Comprehensively profiled social contact patterns in an urban and rural area in Mozambique) | - Age group - Urban/rural |
| Seroprevalence data | Instituto Nacional de Saude, Mozambique^30^ | - Age group - Rural : After waves 2, 3, 4 - Urban : After waves 1, 2, 3 |
| Vaccination rates over time | Instituto Nacional de Saude, Mozambique/ Our World in data^1^ | - Daily - First/second dose |
| Reported cases | Instituto Nacional de Saude, Mozambique/ Our world in data^1^ | - Daily |

*Table 2****.*** *Data sources from Mozambique used to parameterize the transmission model*

#### SI.3.2 Generation of mixing matrix

Our social contact mixing matrix (Fig 2) is derived from contact diaries sampled through age-based quotas in an urban and a rural area during the COVID-19 pandemic between March 2021-March 2022 (N=1242). The socialmixr package^31^ was used to generate symmetric age-specific contact matrices, separately for urban and rural areas. Since contact age groups were estimated with an upper and lower bound, we used 1000 bootstrapped samples to compute the number of contacts between each age group (0-17 years, 18-49 years, 50 years and above). We generated weights by calculating the sampling probabilities for the participant age groups used to develop the sampling frame of the survey and applied these weights during the bootstrapping. Urban and rural daily travel probabilities were estimated for the extent of contact between urban and rural populations based on a travel survey conducted in Mozambique between 2021-2022. Briefly, the survey asked participants from Mahnica (rural) on the frequency of travel to an urban area over the two weeks prior to survey. This frequency was then converted to a daily probability of travel to an urban area among rural participants. Similarly, the survey asked participants from Maputo (urban) on the frequency of travel to a rural area over the two weeks prior to survey, which was then converted to a daily probability of travel to a rural area among urban participants. We then applied these probabilities evenly across the age distribution for who-acquired-infection-from-whom (WAIFW) matrix stratified by both age group and by urban/rural (Fig 2).

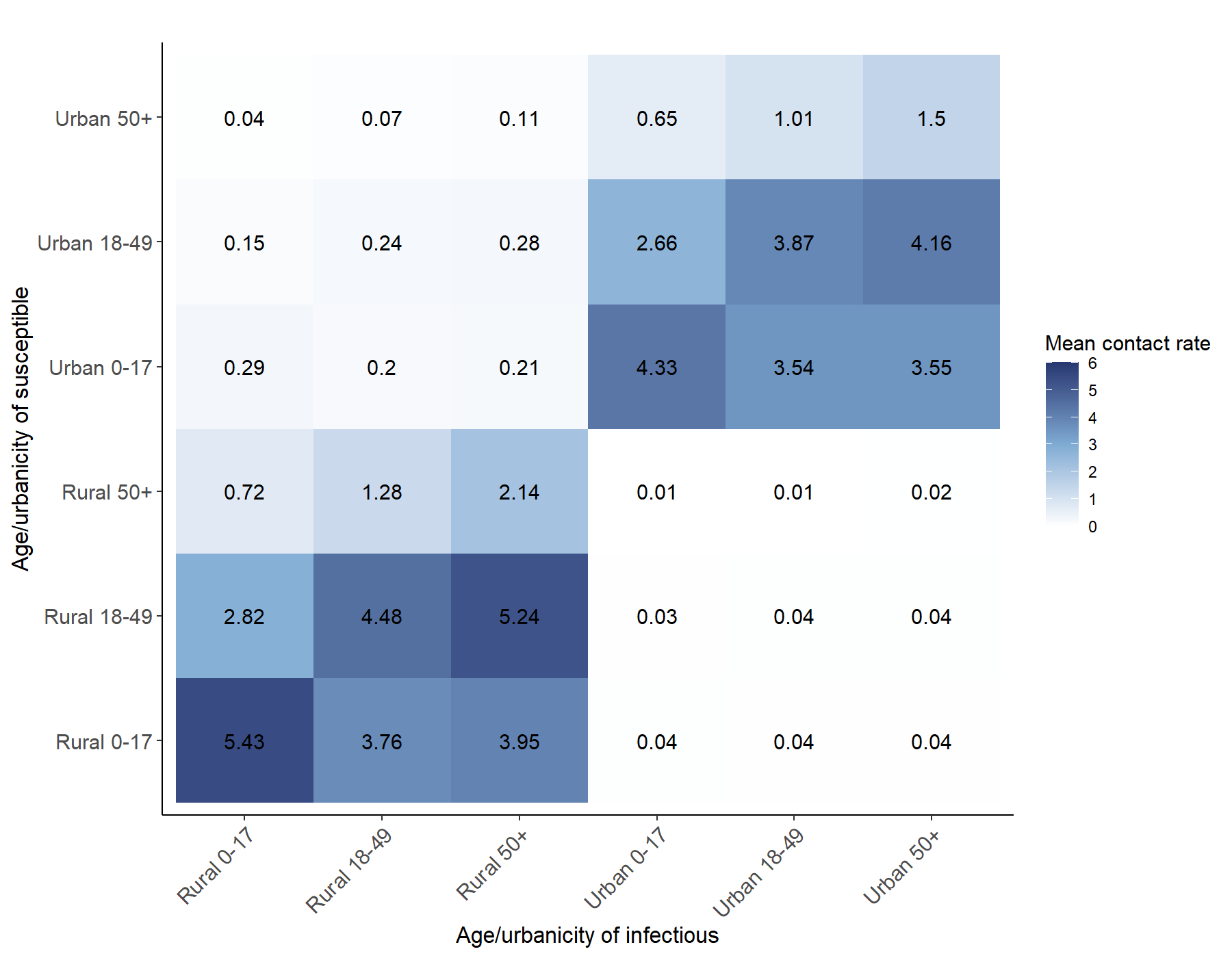

*Figure 2. Matrix of mixing, or who-acquired-infection-from-whom (WAIFW) stratified by age group and by urban/rural. Data used to inform this matrix was collected from an urban and a rural area during the COVID-19 pandemic between March 2021-March 2022.*

#### SI.3.3 Seroprevalence estimates

Several seroprevalence studies have been conducted in Mozambique since the beginning of the COVID-19 pandemic. Between June-Dec 2020 after the first and second waves, the Insituto Nacional de Saude (INS) conducted seroprevalence surveys in 13 urban or peri-urban areas (at least one per province) using the Panbio COVID-19 IgG/IgM rapid test. Surveys were stratified sampled by age group and were intended to be representative of the sampled area. A total of 49,103 individuals were sampled and the area-specific seroprevalence ranged from 0.7% to 7.4%. We pooled these estimates using a simple average to represent the seroprevalence before the start of the second wave in December 2020. Between Dec 2020- Dec 2021, longitudinal seroprevalence surveys using Dried Blood Samples (DBS) were conducted among 2400 individuals in an urban area which forms our seroprevalence estimates for urban areas after the second (winter 2020-2021) and third (summer 2021) COVID-19 waves. Between May 2021 and June 2022, four cross-sectional seroprevalence surveys were conducted in a rural area forms our seroprevalence estimates for rural areas after the second, third and fourth (winter 2021-2022) COVID-19 waves (N= between 666-974). The primary assay used was an S-protein target-specific Luminex assay for IgG and IgM (92% sensitivity and 100% specificity). The most recent Luminex seroprevalence results from February 2022 estimated an overall seroprevalence of 79% in the rural area. Children who are largely unvaccinated have a seroprevalence of 64%, while adults and older adults with high primary series vaccination coverage have a seroprevalence of 86% and 79%, respectively.

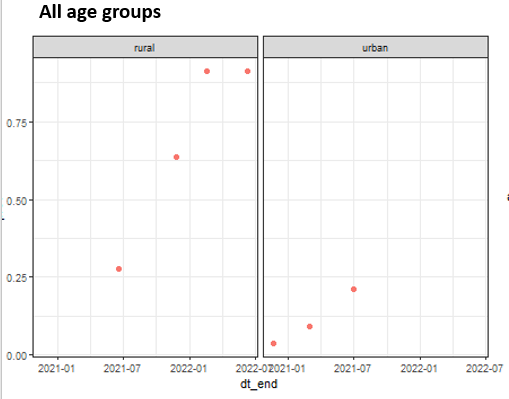

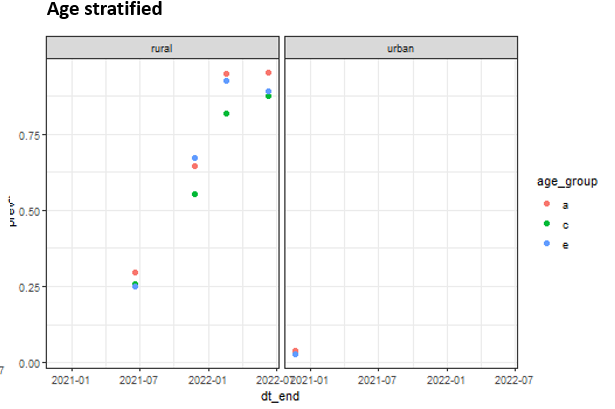

*Figure 3. Seroprevalence point estimates sampled during the COVID-19 pandemic stratified by age group and by urban/rural.*

#### SI.3.4 Vaccination and case data

A publicly-available data source (Our World in Data) and data compiled by Mozambique’s Instituto Nacional de Saude were used to derive reported cases to inform model calibration and the number of first and second doses administered on a daily basis to model the vaccination rate. The latter was converted into a daily vaccination rate for first and second dose.

### SI.4 Calibration results

*SI.4.1 Modeled seroprevalence from the top performing calibration runs*

| **Date** | **Epidemic Period** | **Urban/Rural** | **Age group** | **Modeled median** | **Lower** | **Upper** | **Population samples** |
| --- | --- | --- | --- | --- | --- | --- | --- |
| 10/22/2020 | Post wave1 | Urban | All ages | 1.6% | 1.3% | 2.4% | 3.6% |
|  |  |  | Children | 1.5% | 1.2% | 2.3% | 2.6% |
|  |  |  | Adults | 1.8% | 1.4% | 2.7% | 3.9% |
|  |  |  | Adults >50 yrs | 1.1% | 0.9% | 1.6% | 2.9% |
| 1/30/2021 | Wave 2 | Urban | All ages | 7.9% | 6.8% | 10.0% | 9.0% |
| 5/19/2021 | Post wave 2 | Rural | All ages | 27.8% | 26.2% | 30.6% | 27.6% |
|  |  |  | Children | 25.3% | 22.5% | 28.5% | 25.6% |
|  |  |  | Adults | 31.8% | 29.1% | 35.1% | 29.7% |
|  |  |  | Adults >50 yrs | 25.3% | 23.7% | 27.3% | 25.0% |
|  |  | Urban | All ages | 16.9% | 16.0% | 19.1% | 21.0% |
| 10/1/2021 | Post wave 3 | Rural | All ages | 54.2% | 51.2% | 58.8% | 54.2% |
|  |  |  | Children | 50.8% | 47.0% | 56.1% | 45.2% |
|  |  |  | Adults | 59.2% | 55.7% | 64.0% | 57.2% |
|  |  |  | Adults >50 yrs | 51.2% | 48.9% | 56.9% | 55.4% |
| 1/1/2022 | Post wave 4 | Rural | All ages | 74.7% | 72.0% | 78.1% | 78.6% |
|  |  |  | Children | 65.4% | 61.3% | 70.9% | 63.6% |
|  |  |  | Adults | 86.4% | 84.8% | 88.0% | 86.2% |
|  |  |  | Adults >50 yrs | 82.3% | 81.1% | 84.5% | 78.7% |

*Table 3. Modeled seroprevalence from the top performing calibration runs compared to the population samples of seroprevalence estimates*

#### SI.4.2 Values from top performing calibration runs for calibrated parameters

| **Parameter** | **Initial range** | **Median calibrated value (range)** |
| --- | --- | --- |
| $\beta_{c}$ (relative to $\beta_{e}$) | 0.4-1.2 | 0.358 (0.350-0.370) |
| $\beta_{a}$(relative to $\beta_{e}$) | 0.4-1.2 | 0.479 (0.439-0.528) |
| $\beta_{e}$ | 0.04-0.08 | 0.0747 (0.0712-0.0769) |
| R_0_ | 1.8-2.5 | 2.11 (2.08-2.17) |
| Rate of antibody waning after first infection | 1/400-1/650 | 1/600 (1/500-1/650) |
| Increased transmissibility relative to original strain (delta) | 1.4-1.7 | 1.57 (1.49-1.65) |
| Immune escape (delta) | 1-1.5 | 1.27 (1.10-1.45) |
| Increased transmissibility relative to original strain (omicron) | 2.5-6 | 3.20 (2.93-3.67) |
| Immune escape (omicron) | 1.3-1.9 | 1.55 (1.40-1.80) |

*Table 4. Values from top performing calibration runs for calibrated parameters with median and range.*

### SI. 5 Vaccination impact results for main analysis

#### SI.5.1 Summary table of vaccine impact results

| **Vaccination scenario** | **NNT (older adults)** | **Deaths Averted among older adults** | **Median percent reduction in deaths** | **Deaths among older adults** | **NNT (All ages)** | **Deaths Averted (All ages)** | **Deaths (All ages)** | **No. of campaigns** | **Time of first vaccination (days since start)** |
| --- | --- | --- | --- | --- | --- | --- | --- | --- | --- |
| No vax | - | - | 0% | 6281 (5860-6722) | - | - | 8605 (8026-9218) | - | - |
| Annual | 1941 (1805-2112) | 3284 (3019-3531) | 52% | 2999 (2703-3315) | 1571 (1443-1735) | 4058 (3675-4417) | 4548 (4095-5035) | 10 (10-10) | 300 (300-300) |
| Biennual | 1443 (1223-1733) | 2206 (1837-2601) | 35% | 4059 (3646-4579) | 1173 (967-1445) | 2713 (2204-3289) | 5865 (5263-6619) | 5 (5-5) | 300 (300-300) |
| 50% thresh | 1499 (1252-1905) | 1274 (1002-1526) | 20% | 5017 (4504-5507) | 1124 (930-1429) | 1700 (1336-2055) | 6918 (6185-7587) | 3 (3-3) | 1667 (1596-1782) |
| 55% thresh | 1499 (1358-1762) | 1699 (1445-1875) | 27% | 4592 (4191-4979) | 1118 (1006-1305) | 2278 (1952-2532) | 6342 (5763-6903) | 4 (4-4) | 1356 (1318-1509) |
| 60% thresh | 1552 (1391-1906) | 2053 (1671-2289) | 33% | 4242 (3890-4654) | 1151 (1028-1393) | 2768 (2287-3099) | 5844 (5358-6444) | 5 (5-5) | 1132 (1011-1242) |
| 65% thresh | 1727 (1572-2348) | 2584 (1900-2838) | 41% | 3742 (3318-4266) | 1327 (1204-1710) | 3363 (2608-3704) | 5259 (4789-5887) | 7 (7-7) | 822 (604-962) |
| 70% thresh | 2093 (1922-2313) | 3047 (2757-3319) | 48% | 3233 (2908-3596) | 1632 (1486-1816) | 3907 (3511-4290) | 4698 (4216-5237) | 10 (10-10) | 436 (423-457) |
| 75% thresh | 2355 (2169-2723) | 3794 (3284-4118) | 60% | 2493 (2164-2991) | 1840 (1697-2107) | 4853 (4254-5263) | 3780 (3319-4375) | 14 (14-14) | 255 (255-256) |
| 80% thresh | 3151 (2943-3429) | 4456 (4095-4771) | 71% | 1835 (1602-2042) | 2456 (2290-2693) | 5719 (5214-6133) | 2909 (2541-3231) | 22 (22-22) | 109 (109-109) |

*Table 5. Summary results on NNT, number of deaths, number of deaths averted, median percent reduction in deaths and vaccination timing and frequency based on different vaccination strategies in the epidemic scenario driven by waning immunity. Quantitative results presented as median and 2.5^th^-97.5^th^ percentile ranges. The median percent reduction in deaths was calculated using the medians of deaths averted and deaths among older adults in a no vaccination scenario*

#### SI.5.2 Distribution of NNT and number of deaths across all age groups

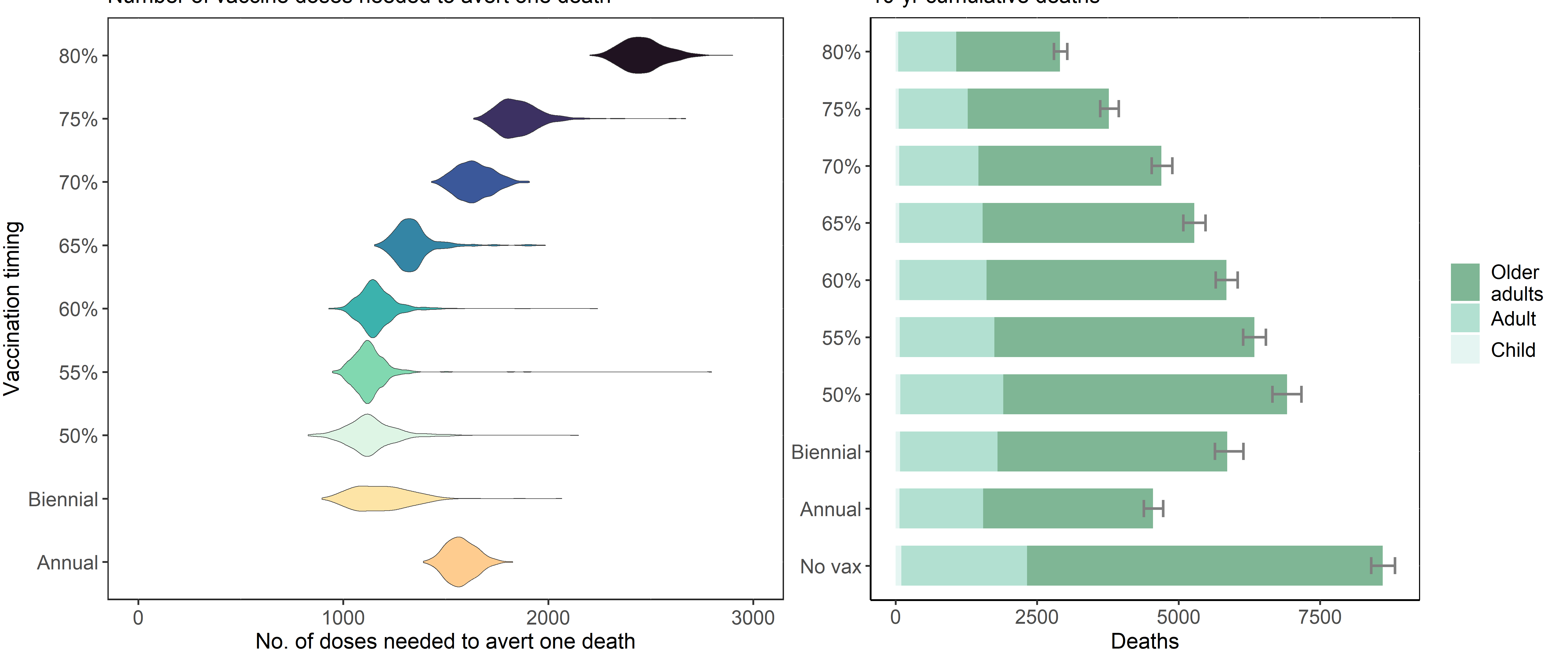

*Figure 4. (Left) Distribution of the number of vaccine doses needed to avert one death among all ages (NNT) by vaccination timing strategy (biennual, annual, triggered based on seroprevalence thresholds of between 50%-805) based on random sampling for annual transmission under an epidemic scenario driven by waning immunity. (Right) Cumulative deaths over ten years by age group (dark green=child, medium shade = adults, lightest shade =adults >50 years). Error bar represents 25^th^-75^th^ percentile of cumulative deaths across all age groups*

#### SI.5.3 Susceptibility landscape over time stratified by age group

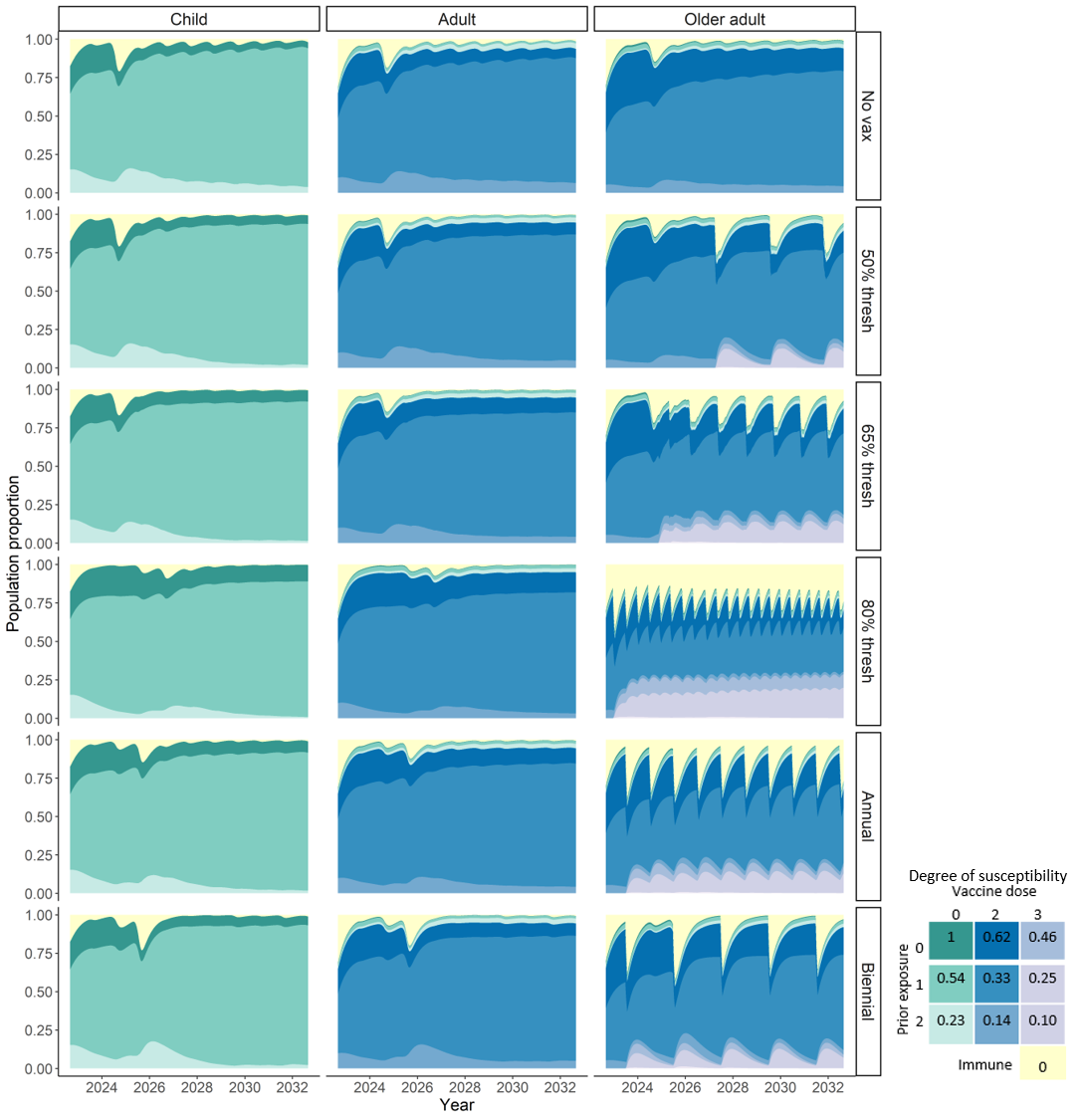

*Figure 5. Colored density represents population proportion within susceptible or immune tiers over time, ranging from fully immune (yellow) to up to 2 prior infections and 3 vaccination doses. The blue and purple densities indicate proportion of individuals in the 2- and 3- vaccine dose susceptibility tiers, respectively. The darkest shades within each color have no prior infection and are the most susceptible, with lighter shades indicating more exposure and decreased susceptibility. Individuals can wane from the 3-dose susceptibility tier to the 2-dose tier and from the 3-prior infection tier to the 2-prior infections at a rate of 1/365 days. The degree of susceptibility is indicated in the grid legend and is relative to totally susceptible, with 1 indicating fully susceptible and 0 fully immune. In scenarios with vaccination, campaigns generate spikes in the proportion of individuals fully immune (yellow). More frequent vaccination campaigns result in higher proportion immune (yellow) and longer period in the compartment with highest protection (purple).*

#### SI.5.4 Correlations between seroprevalence, susceptibility and cumulative deaths

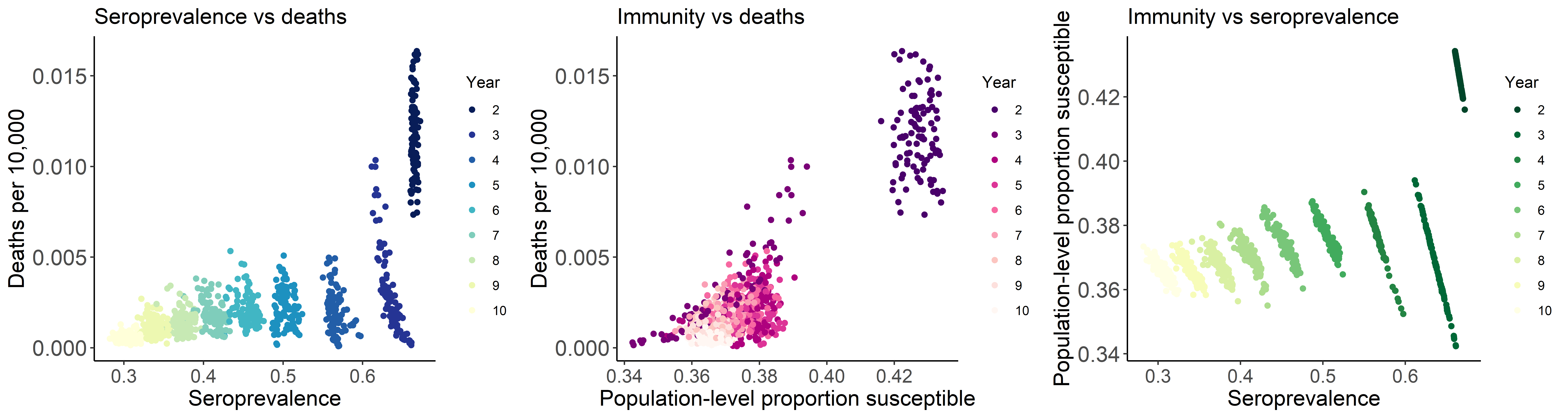

*Figure 6. (Bottom) Scatter plots under no vaccination for: (Left) seroprevalence of older adults prior to each wave and deaths per 10,000; (Middle) population-level proportion susceptible of older adults prior to each wave and deaths per 10,000; (Right) seroprevalence of older adults prior to each wave and the population-level proportion susceptible of adults prior to each wave.*

We had hypothesized that serology-triggered strategies to target time periods of greater susceptibility would perform better than fixed interval strategies. When our results did not support this, we explored potential reasons for similar efficiencies between serology-triggered vaccination strategies versus fixed time strategies. We assessed pairwise correlations between seroprevalence and susceptibility prior to each wave, and deaths per 10,000 among older adults (Figure 6) in the base scenario with no vaccination. We expected a negative correlation where lower seroprevalence at the start of each wave is correlated with more deaths. Instead, we find that seroprevalence is positively correlated with deaths (R^2^=0.46). We calculate a summary estimate of susceptibility where the proportion of the population in each susceptible tier is multiplied by the degree of susceptibility, with theoretical ranges from 1 for a completely susceptible population to 0 for a completely immune population. We expect a positive correlation where increased susceptibility is correlated with increased deaths and we find a positive correlation with R^2^=0.80. Lastly, we assess the correlation between susceptibility and seroprevalence. We expect a negative correlation where increased seroprevalence is associated with decreased susceptibility. We find the association within a year to be negative but positive overall over the entire 10-year simulation period, driven by comparatively high seroprevalence and high susceptibility in year 2. The lack of protection against infection among children drives earlier epidemics to the extent that protection in older adults is a poor marker for large epidemics that result in higher population-level deaths.

### SI.6 Sensitivity analysis using a randomly-timed epidemic scenario

#### SI.6.1 Descriptive results

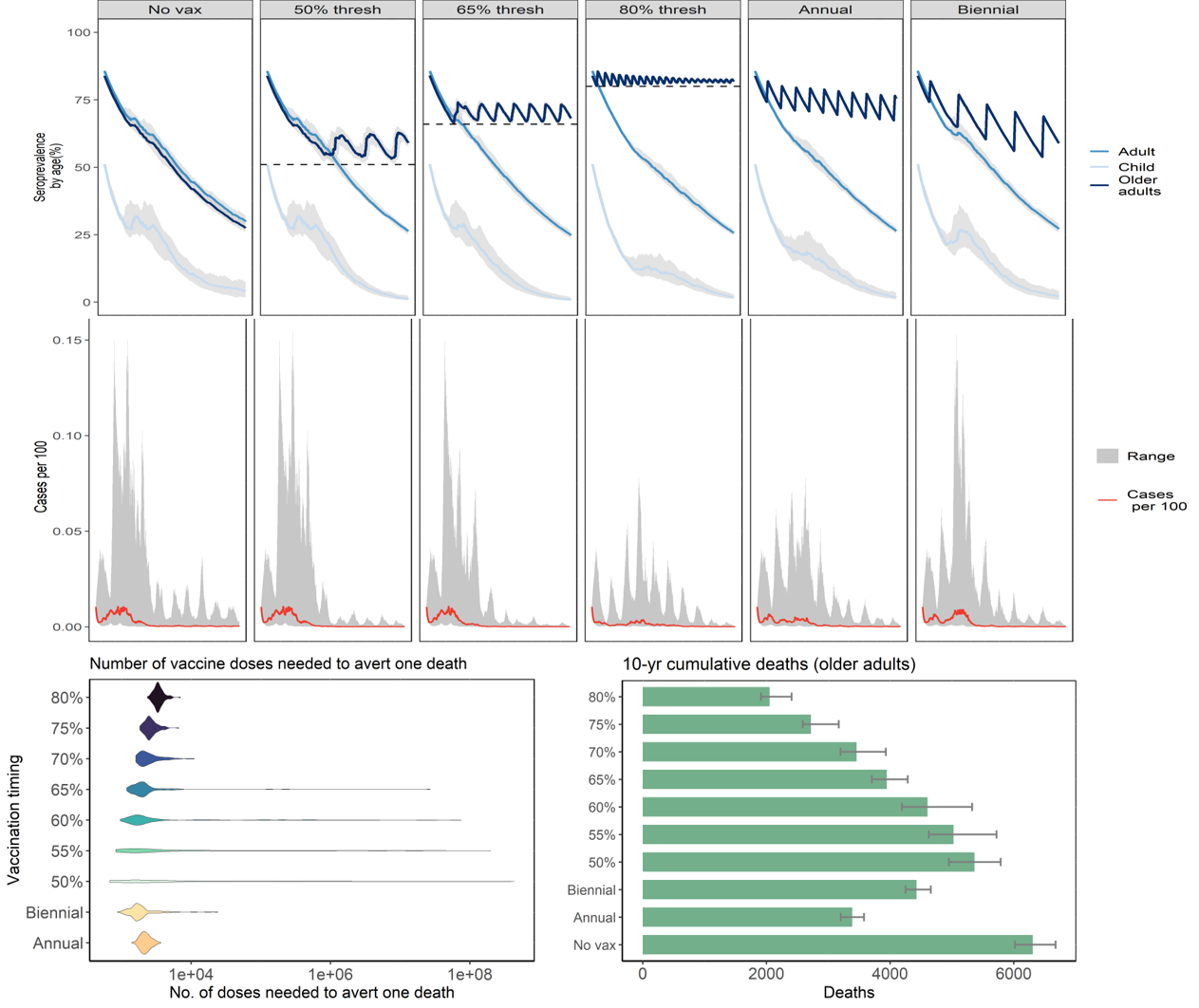

*Figure 7. (Top row) Modeled 10-year seroprevalence (gray = 2.5th-97.5th percentile) over time for children (lightest blue), adults (medium blue) and older adults >50 years (darkest blue) under 1) no additional vaccinations; 2) vaccinations timed by seroprevalence trigger thresholds of 50%, 65% and 80% and 3) vaccinations timed annually and biennially. (Middle row) Modeled 10-year cases per 100 individuals over vaccination scenarios (gray shading=ranges from random Rt sampling, red= median); (Left) Distribution of the number of vaccine doses needed to avert one death among all ages (NNT) by vaccination timing strategy (biennual, annual, triggered based on seroprevalence thresholds of between 50%-80%) based on a randomly-timed epidemic scenario driven by immune waning (Right) Cumulative deaths over ten years by age group (dark green=child, medium shade = adults, lightest shade =adults >50 years). Error bar represents 25^th^-75^th^ percentile of cumulative deaths across all age groups*

#### SI.6.2 Summary table of vaccine impact results

| **Vaccination scenario** | **NNT (older adults)** | **Deaths Averted among older adults** | **Deaths among older adults** | **NNT (All ages)** | **Deaths Averted (All ages)** | **Deaths (All ages)** | **No. of campaigns** | **Time of first vaccination (days since start)** |
| --- | --- | --- | --- | --- | --- | --- | --- | --- |
| Annual | 2190 (1679-3209) | 2910 (1987-3798) | 3389 (2958-4190) | 1741 (1310-2853) | 3663 (2238-4869) | 4963 (4326-6242) | 10 (10-10) | 300 (300-300) |
| Biennial | 1695 (1067-8841) | 1879 (390-2988) | 4424 (3946-5967) | 1362 (814-5478) | 2295 (398-3810) | 6272 (5626-8409) | 5 (5-5) | 300 (300-300) |
| 50% thresh | 1567 (760-33131) | 1220 (58-2517) | 5162 (4409-6014) | 1175 (562-24122) | 1626 (79-3403) | 7059 (6022-8203) | 3 (3-3) | 1696 (1484-1942) |
| 55% thresh | 1712 (880-21798) | 1488 (118-2895) | 4878 (4176-6003) | 1270 (643-15841) | 2006 (163-3965) | 6697 (5700-8188) | 4 (4-4) | 1402 (1230-1678) |
| 60% thresh | 1793 (1080-14704) | 1803 (217-3092) | 4504 (3899-5857) | 1349 (793-10698) | 2441 (298-4214) | 6268 (5348-8016) | 5 (5-6) | 1051 (867-1352) |
| 65% thresh | 1971 (1292-5338) | 2263 (835-3452) | 3931 (3358-5701) | 1527 (954-4554) | 2920 (979-4677) | 5467 (4840-7894) | 7 (7-7) | 701 (619-928) |
| 70% thresh | 2261 (1634-6429) | 2819 (893-3904) | 3460 (2812-5546) | 1732 (1275-6000) | 3679 (960-5003) | 4970 (4141-7826) | 10 (9-10) | 457 (429-694) |
| 75% thresh | 2537 (1948-4233) | 3520 (2049-4584) | 2719 (2353-4528) | 1989 (1508-3682) | 4488 (2360-5923) | 4047 (3531-6508) | 14 (13-14) | 270 (255-303) |
| 80% thresh | 3376 (2617-5103) | 4158 (2586-5367) | 2053 (1703-3929) | 2618 (2019-4255) | 5357 (3088-6954) | 3206 (2650-5827) | 22 (20-22) | 110 (109-139) |

*Table 6. Summary results on NNT, number of deaths, number of deaths averted and vaccination timing and frequency based on different vaccination strategies in the epidemic scenario driven by randomly-timed annual epidemic and by waning immunity. Quantitative results presented as median and 2.5th-97.5th percentile ranges.*

### SI. 7 Results from the 10-year epidemic trajectory driven by immune escape

#### SI.7.1 Descriptive results

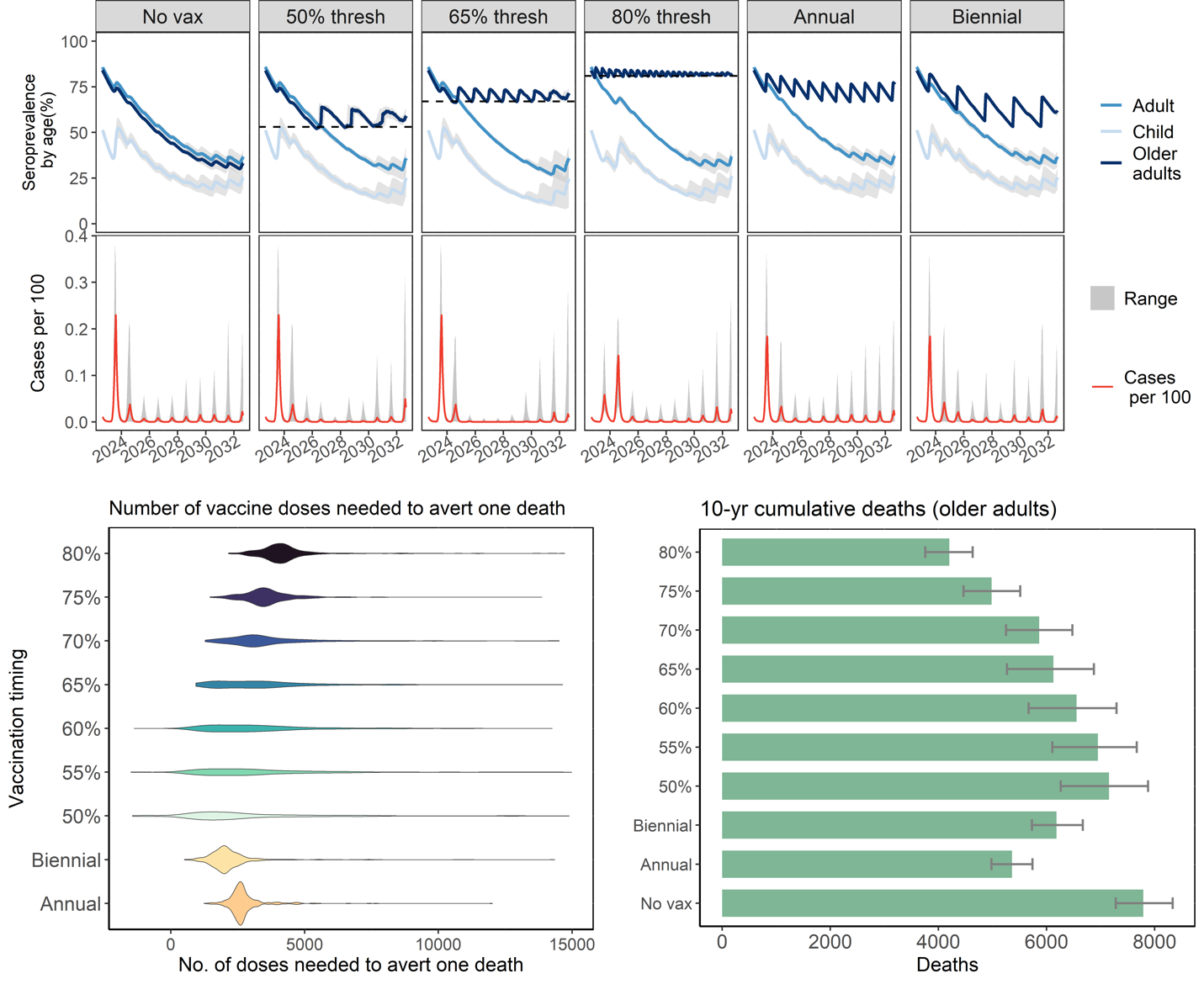

*Figure 8. (Top) Modeled 10-year seroprevalence (gray = ranges) over time using a high immune escape scenario for children (lightest blue), adults (medium blue) and older adults >50 years (darkest blue) under 1) no additional vaccinations; 2) vaccinations timed by seroprevalence trigger thresholds of 50%, 65% and 80% and 3) vaccinations timed annually and biennially. (Middle) Modeled 10-year cases per 100 over vaccination scenarios (gray=ranges from random R0 sampling, red= median); (*Bottom left) *Distribution of the number of vaccine doses needed to avert one death (NNT) by vaccination timing strategy (biennial, annual, triggered based on seroprevalence thresholds of between 50%-80%); (Bottom right) Cumulative deaths over ten years among older adults. Error bar represents 25^th^-75^th^ percentile of cumulative deaths for older adults.*

In the base case high immune escape scenario with no additional vaccinations, multiple peaks arise that is more intense in the first year (Figure XXX). Similar to the high waning immunity scenario, the seroprevalence declines over time and increases in response to surges in cases. Across model runs, the median cumulative number of deaths over 10 years is 10558 (25th-75th percentile: 9885-11282) for all ages, 141 (25th-75th percentile: 132-150) for children, 2628 (25th-75th percentile: 2468-2804) for adults and 7791 (25th-75th percentile: 7281-8334) for older adults.

With seroprevalence-informed vaccination triggers, a similar number of campaigns are triggered compared to the high waning immunity scenario. Compared to a median of 7,791 deaths among older adults over ten years in a scenario with no additional vaccination, vaccinating each time the seroprevalence among older adult falls below 50% and 80% results in a median of 7,157 and 4,200 deaths respectively. Among the vaccination strategies guided by seroprevalence, the median number needed to vaccinate to avert one death (NNT) reaches a minimum at a 50% threshold where 3 campaigns providing a total of 1.9 million vaccine doses result in a median of 1,526 fewer deaths and a median NNT of 452. The NNT increases for higher seroprevalence thresholds with an NNT of 4,092 for an 80% threshold. In comparison, annual and biennial vaccination of older adults results in a median of 5,363 and 6,186 deaths among older adults, respectively and NNTs of 2,617 and 2,013, respectively. Similar patterns are observed for the NNT across all age groups (number needed to vaccinate to avert one death in the entire population).

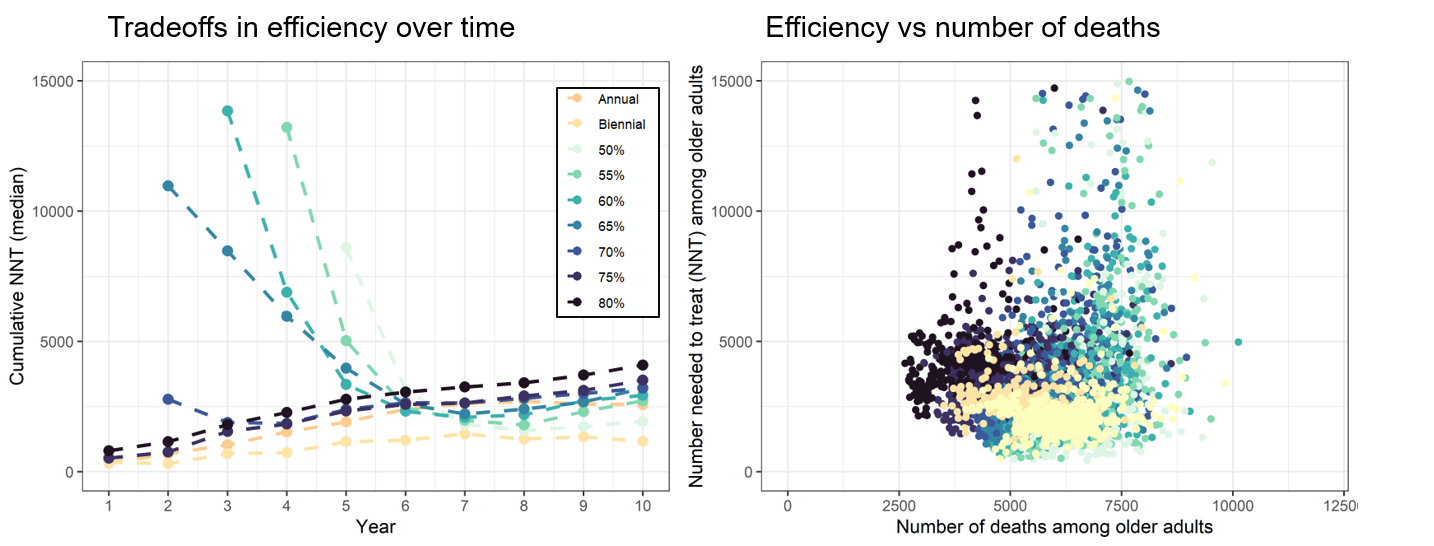

*Figure 9. (Left) Efficiency over time estimated by cumulative NNT for the epidemic scenario driven by immune escape. (Right) Scatterplot of efficiency (NNT) versus number of deaths for the epidemic scenario driven by immune escape.*

#### SI.7.2 Summary table of vaccine impact results

| **Vaccination scenario** | **NNT (older adults)** | **Deaths Averted among older adults** | **Deaths among older adults** | **NNT (All ages)** | **Deaths Averted (All ages)** | **Deaths (All ages)** | **No. of campaigns** | **Time of first vaccination (days since start)** |
| --- | --- | --- | --- | --- | --- | --- | --- | --- |
| No vax | - | - | 7791 (7281-8334) | - | - | 10558 (9885-11282) | - | - |
| Annual | 2617 (2428-2827) | 2432 (2242-2616) | 5363 (4980-5739) | 2405 (2239-2620) | 2638 (2405-2831) | 7941 (7382-8495) | 10 (10-10) | 300 (300-300) |
| Biennual | 2013 (1718-2392) | 1556 (1298-1802) | 6186 (5731-6670) | 1905 (1555-2273) | 1635 (1340-1946) | 8853 (8224-9555) | 5 (5-5) | 300 (300-300) |
| 50% thresh | 2138 (676-4678) | 452 (0-1083) | 7157 (6261-7878) | 1439 (369-3848) | 535 (0-1373) | 9773 (8571-10774) | 3 (3-3) | 1420 (1353-1515) |
| 55% thresh | 2728 (1145-5414) | 675 (82-1232) | 6949 (6109-7670) | 2106 (793-4479) | 798 (30-1543) | 9563 (8378-10519) | 4 (4-4) | 1180 (1137-1249) |
| 60% thresh | 2931 (1550-5072) | 1004 (388-1780) | 6557 (5670-7294) | 2368 (1110-4090) | 1157 (363-2299) | 9105 (7772-10070) | 6 (5-6) | 945 (920-995) |
| 65% thresh | 3198 (1883-4852) | 1423 (865-2341) | 6128 (5266-6878) | 2667 (1380-4244) | 1650 (890-3012) | 8524 (7229-9597) | 8 (7-8) | 762 (732-808) |
| 70% thresh | 3217 (2545-4471) | 1900 (1251-2328) | 5865 (5249-6480) | 2697 (1944-3610) | 2256 (1472-2870) | 8314 (7386-9100) | 10 (10-11) | 536 (500-579) |
| 75% thresh | 3514 (3083-4002) | 2697 (2355-3075) | 4983 (4464-5514) | 3109 (2655-3598) | 3031 (2610-3525) | 7380 (6593-8162) | 15 (15-15) | 215 (215-215) |
| 80% thresh | 4092 (3699-4507) | 3546 (3228-3909) | 4200 (3755-4633) | 3637 (3227-4075) | 3971 (3577-4464) | 6514 (5860-7155) | 23 (22-23) | 92 (92-92) |

*Table 7. Summary results on NNT, number of deaths, number of deaths averted (for older adults and all ages) and vaccination timing and frequency based on different vaccination timing strategies under an epidemic scenario driven by immune escape*

#### SI.7.3 Distribution of NNT and number of deaths across all age groups

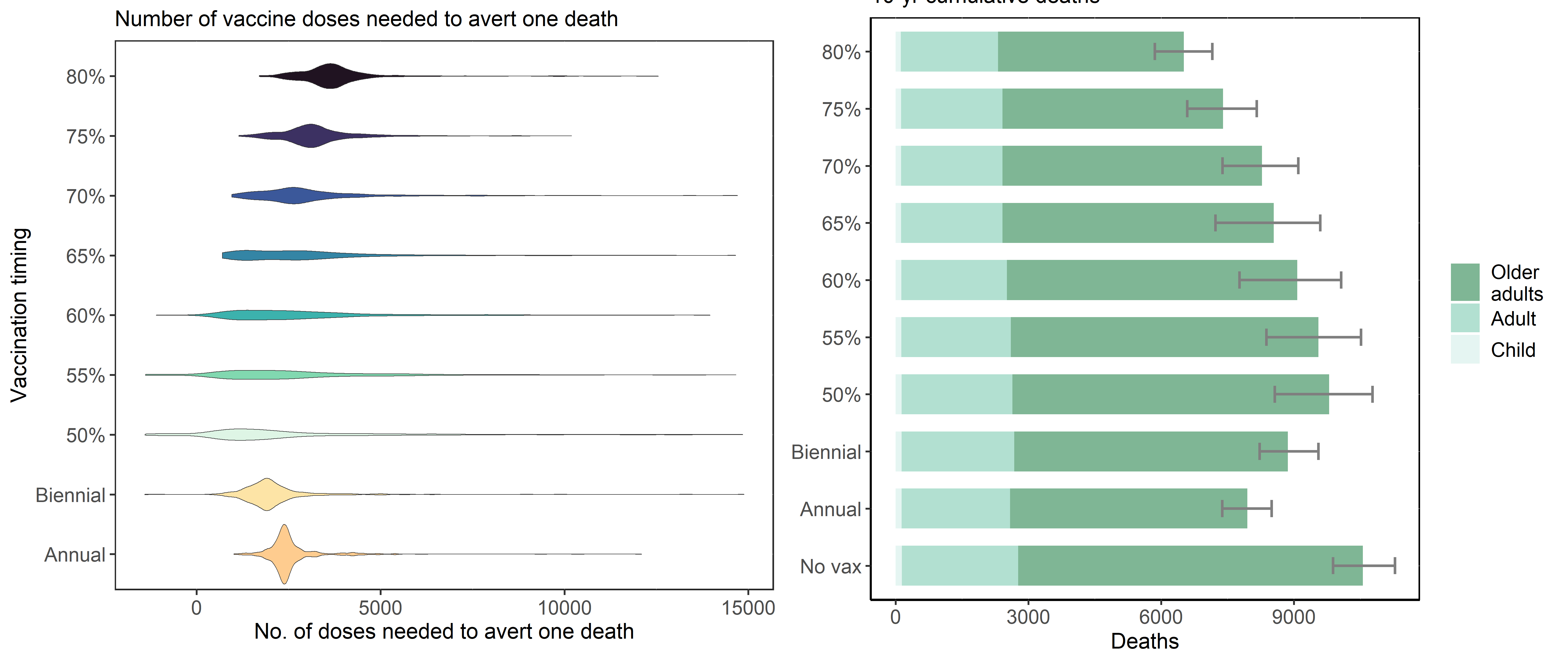

*Figure 10. (Left) Distribution of the number of vaccine doses needed to avert one death among all ages (NNT) by vaccination timing strategy (biennual, annual, triggered based on seroprevalence thresholds of between 50%-80%) based on random sampling for annual transmission under an epidemic scenario driven by immune escape. (Right) Cumulative deaths over ten years by age group (dark green=child, medium shade = adults, lightest shade =adults >50 years). Error bar represents 25^th^-75^th^ percentile of cumulative deaths across all age groups*

#### SI.7.4 Susceptibility landscape over time stratified by age group

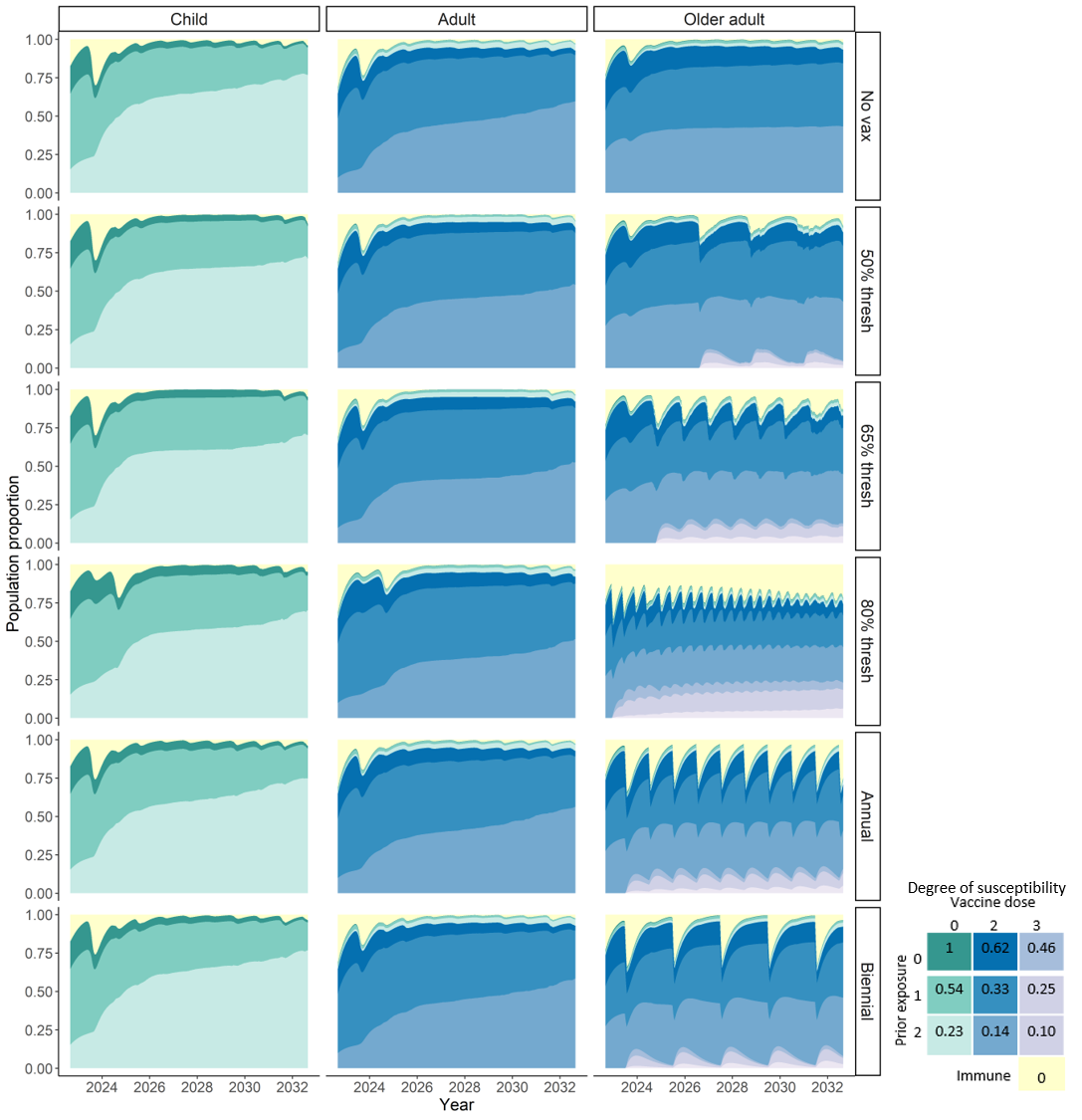

*Figure 11. Colored density represents population proportion within susceptible or immune tiers over time, ranging from fully immune (yellow) to up to 2 prior infections and 3 vaccination doses. The blue and purple densities indicate proportion of individuals in the 2- and 3- vaccine dose susceptibility tiers, respectively. The darkest shades within each color have no prior infection and are the most susceptible, with lighter shades indicating more exposure and decreased susceptibility. Individuals can wane from the 3-dose susceptibility tier to the 2-dose tier and from the 3-prior infection tier to the 2-prior infections at a rate of 1/365 days. The degree of susceptibility is indicated in the grid legend and is relative to totally susceptible, with 1 indicating fully susceptible and 0 fully immune. In scenarios with vaccination, campaigns generate spikes in the proportion of individuals fully immune (yellow). More frequent vaccination campaigns result in higher proportion immune (yellow) and longer period in the compartment with highest protection (purple).*

### SI.8 Summary of literature review of key parameters

#### SI.8.1 Evidence on seroconversion after infection or vaccination

| **Reference** | **Evidence** | **Location** | **Study design and population** |
| --- | --- | --- | --- |
| **Post-infection** | | | |
| Lou et al^32^ | Seroconversion rates for total antibody, IgM and IgG were 98.8%, 93.8% and 93.8%, respectively | China | Longitudinal (N=80) among PCR confirmed |
| Oved et al^19^ | Using multiple assays, antigen targets, estimated 5% of PCR-infected individuals remained persistently seronegative | Israel | Longitudinal (698) among PCR positive individuals |
| Van Elslande et al^33^ | •22% of mild cases and 2.6% of severe cases never seroconverted | Belgium | Paired sera (N=236) PCR confirmed from hospitals |
| **Post-vaccination** | | | |
| Ali et al^34^ | Post-vaccination antibody-levels are higher among individuals with prior infection (30-40% higher) | Kuwait | N=1025 among vaccinated individuals |
| Anichini et al^35^ | •Similar post-vax IgG levels between individuals with and without prior infection •Higher neutralizing titers among those with prior infection | Italy | N=100 HCWs post first dose vax, 38 with history of infection |
| Assis et al^36^ | •Post-vax, previously infected individuals developed higher antibody titers to vaccine than non pre-exposed individuals •Higher antibody levels among those with severe disease | California, US | N=8761 before and after vax campaign |
| Ward et al^37^ | •After single dose Pfizer after 21 days, 84% of people under 60 years tested positive and 90% among those with prior infection (across all age groups) •After two dose Pfizer, ~100% test positive •After two dose AZ, <90% for those aged 35 and above and 73% among oldest age group | UK | REACT study 155,172 with valid IgG results |

#### SI.8.2 Evidence for durability of antibody

| **Reference** | **Evidence** | **Location** | **Study design and population** |
| --- | --- | --- | --- |
| CDC | Antibodies derived from infection last between 3-6 months and up to 11 months |  |  |
| He et al^38^ | 90% positive for IgGs 6-8 months after seroconversion | Wuhan, China | Longitudinal, among those positive for IgGs |
| Alfego et al^39^ | •max of 90% seropositive for IgG (S and N-proteins) 21 days post-index •N-protein seropositivity declined to 68.2% through 293 days;  •S-antibody seropositivity declined to 88% through 300 days •Age associated with seroreversion, ~seropos approx 15% lower in >-65 years after 280 days | US | Cross-sectional (N=39,086) individuals with PCR-confirmed infection between Mar 2020-Jan 2021 |
| Peluso et al^40^ | •Time to seroreversion ranged by assay, ranging from 96 days for N(frag) – Lum to 925 days for S-DiaSorin. S-Lum had mean time to sero-reversion of 400-500 days among the non-hospitalized •Lower antibody titers among individuals with mild infection | California, US | Longitudinal (n=128) |
| Wei et al^41^ | • Anti-spike IgG half-life was 184 days • Ab levels associated with protection against reinfection likely last 1.5-2 years on average with levels associated with protection from severe infection present for several years | China | Among PCR positive individuals |
| Wang et al^42^ | positivity rates for IgM, IgG, anti-RBD IgG, and NAb fell to 20.4% (39/191), 97.9% (187/191), 97.4% (186/191), and 95.8% (183/191), respectively, during 9–10 months post symptom onset | China | 215 individuals established in Feb 2020 |
| Harris et al^43^ | •Predicted proporstions sero-reverting after 52 weeks were 100% for Abbot, 59% Euroimmun, 41% RBD, 10% Roche (N), <2% Roche (S) | UK | Longitudinal (N=264) among those seropositive for >2 assays |
| Shioda et al^44^ | Time from seroconversion to seroreversion was 3-4 months | NYC and Connecticut | Cross-sectional serology data (N=1800 for each site, each cross) |
| Swartz et al^45^ | Model fitting of antibody response suggests that individuals may remain antibody positive from natural infection beyond 500 days | Texas, US | Longitudinal-ish? (N=4553) among those with at least one antibody test with 1-3 Ab tests over 11 months |
| Yang et al^46^ | •Anti-RBD IgG peaked at 120 days and declined •At 400-480 days, undetectable neutralizing acitivty found in 14% (16/111) of mild •At 330-480 days, 50% (5/10) undetectable among asymptomatic infections | China | Longitudinal (N=214) convalescents without additional exposure after recovery or vaccination |
| Van Elslande et al^33^ | •22% of mild cases and 2.6% of severe cases never seroconverted •Of mild seroconverters, 18.8%, 40% and 61% were seronegative in the windows 60-119 days, 120-179 days and 180-240 days •Of severe seroconverters, number was 1.9%, 10.8% and 29.4% | Belgium | Paired sera (N=236) PCR confirmed from hospitals |
| Rees et al^47^ | Using age-structured reverse catalytic model, estimated antibody persistence lasted between 0.9 (0.6-1.6 years) and 5.8 (2.0-7.4 years) | Multiple countries | Historical seroprevalence data of four circulating HCoVs |
| Feng et al^48^ | Increasing Ab titers following vaccination associated with increasing VE | UK | Cohort of AZ efficacy trial |
| Grandjean et al^49^ | S antibody predicted to remain detectable in 95% of participants until 465 days compared to 75% of N-antibodies (nice figures for reference) | UK | Cohort (N=349) of seropostivie HCWs, data for 200 days |
| Ward et al^37^ | •Between 1-2 dose of pfizer, Ab fell to 64% of peak levels (10-12 weeks) after a peak at 4-5 weeks | UK | REACT study 155,172 with valid IfF results |

#### SI.8.3 Evidence for vaccine effectiveness

| **Reference** | **Evidence** | **Location** | **Study design and population** |
| --- | --- | --- | --- |
| Cromer et al^50^ | Modelling of predicted vaccine efficacy (based on neutralisation titres) against variants over time suggested that protection against symptomatic infection might decrease below 50% within the first year after vaccination | Multiple, pooled data across 24 studies | Modeled based on antibody neutralisation titres |
| Hall et al^51^ | • Two dose Pfizer, VE=85% at 14-73 days and VE = 51% at 201 days •Two dose AZ VE = 49% (16-69%) at 14-73 days •Two dose AZ VE = 47% (26-63%) at 74-133 days •Two dose AZ VE = 51% (18-71%) after 133 days •Infection-acquire immunity in unvaxed participants (<5%) waned after 1 year (dropped from 86% (81-89%) within 1 year to 70% (38-84%) after 1 year.  •Among infected and vaxed 1 dose, VE 90% (60-97%) at >1 year after primary infection •Infected and vaxed 2 dose, VE 95% (82-99%) at >1 year after primary infection | Qatar (Expats) | Test negative case-control (nested in N=35K undergoing aoutine symptomatic screening) |
| Altarawneh et al^52^ | •Pfizer/moderna (6 months prior)+ no prior infection against symptomatic omicron= -1.1% (basically none) •Three dose pfizer/moderna +no prior infection = 52.2% (48-56) •Effectiveness of IE, VE and hybrid against severe, critical, fatal BA.1 infection was >95% •2-dose VE alone against severe/critical fatal BA.2 infection = 77% •3-dose VE against severe/critical/fatal BA.2 infection = 98% | Qatar (Expats) | Matached, test-negative, case-control |
| Murugesan et al^53^ | •VE (AZ) (vaxed Jan 2021) with no prior infection =32% (24-39%) | South India | Cohort study, infection outcomes during delta wave (April 2021) |
| Poukka et al^54^ | •VE (AZ) against infection from delta 14-90 days = 88% (71-95) •VE (AZ) against infection from delta91-180 days = 62% (17-95) •VE(AZ) against severe disease from delta 14-90 days = 100 (25-100%) •VE (AZ) against severe disease from delta 91-180 days = 81 (9-96%) | Finland | Retrospective cohort study |
| Nordstrom et al^55^ | •VE (AZ) against symptomatic disease 31-120 days = 45% (28-60%) •VE (AZ) against symptomatic disease >120 days = 19% (-97-28%) | Sweden | Retrospective cohort study |
| Pattni et al^56^ | •VE (AZ) one dose in reducing susceptibility to infection = 39%(34-43%) for delta •VE (AZ) two dose in reducing susceptibility to infection = 64% (61-67%) for delta | UK | Retrospective cohort from anonymized public health data linked to PCR data |
| Tan et al | •VE (mRNA) two dose against infection with Delta = 45% (40-50%), against Omicron = 21% (7-34%) •VE (mRNA) for booster against infection was 44% (38-50%) for delta and 40% (35-40%) for omicron •VE against severe disease by booster for omicron was 83% (76-88%), VE against severe disaese by primary series for delta was 80% (73-85%) See figure image | Singapore | Test-negative case-control study |

#### SI.8.4 Evidence for effectiveness of prior infection against subsequent infection

| **Reference** | **Evidence** | **Location** | **Study design and population** |
| --- | --- | --- | --- |
| Altarawneh et al^52^ | •IE alone against symptomatic BA.2 infection = 46.1% (40-52%) •IE + two dose pfizer/moderna = 55% (51-59) •IE + three dose pfizer/moderna = 77% (72-81%) •IE alone against severe/critical fatal BA.2 infection = 73% •Hybrid against severe/critical/fatal BA.2 infection = 98%-100% | Qatar (Expats) | Matached, test-negative, case-control |
| Kojima et al^57^ | •Weighted average = 90.4% (range 82-100) up to 10 months fololow up | Multiple HICs | Systematic review (10 articles), published before June 2021 |
| Murugesan et al^53^ | •IE against symptomatic infection = 86% (77-92%) •IE+vax in Jan 2021 = 91% (84-95%) | South India | Cohort study, infection outcomes during delta wave (April 2021) |
| Hansen et al^58^ | •Protection against repeat infection = 81% (75-85%) | Denmark | Compared reinfection in second surge (Sep-Dec 2020) between individuals with positive and negative PCR tests during first surge |
| Lumley et al^59^ | •Protection against re-infection among Ab positive = 89% (56-97%) (up to 5 months) | UK | Cohort study, seropositive and negative HCWs beginning April 2020 |
| Letizia et al^60^ | •Protection against re-infection among Ab positive = 92% (72-89%) •Higher baseline Ab titres associated with decreased risk of re-infection | US | Cohort of marine recruits (N=3168) followed for 6 weeks |
